# Gut microbiota-derived lithocholic acid leads to intestinal barrier dysfunction via LPCAT1 in irritable bowel syndrome

**DOI:** 10.1101/2025.01.13.25320445

**Authors:** Lixiang Zhai, Baohua Li, Shujun Xu, Jiao Peng, Yingdi Zhang, Jiayu He, Gengyu Bao, Yunlyu Li, Huan Deng, Ling Zhao, Ziwan Ning, Junfang Lyu, Chengyuan Lin, Hoi Leong Xavier Wong, Zhaoxiang Bian, Haitao Xiao

**Author notes:** **Correspondence:** Lixiang Zhai, Ph.D.; Zhaoxiang Bian, MD, Ph.D.; Haitao Xiao, Ph.D. These authors contribute equally to this work (Lixiang Zhai, Baohua Li, Shujun Xu, Jiao Peng). **Declaration of interests** The authors declare that they have no competing interests.

## Abstract

Intestinal barrier dysfunction is widely observed in patients with irritable bowel syndrome (IBS) and significantly contributes to the persistence of IBS symptoms. The association between gut dysbiosis and the pathogenesis of IBS, as well as its connection to intestinal barrier dysfunction, has been established. However, the precise roles of gut bacteria in inducing intestinal barrier dysfunction and the underlying mechanisms of IBS remain unclear. In the present study, we showed that microbiota-derived lithocholic acid (LCA) is positively associated with intestinal barrier dysfunction biomarkers in patients with IBS-D. We found excessive LCA disrupts intestinal barrier function in normal mice and aggravates colonic inflammation in a mouse model of experimental colitis. Mechanistically, LCA modulates phospholipid metabolism and compromises the intestinal barrier by directly activating lysophosphatidylcholine acyltransferase 1 (LPCAT1). Our results demonstrated that LCA significantly upregulates LPCAT1 expression, and its overexpression leads to intestinal barrier dysfunction and promotes colonic inflammation via the activation of matrix metalloproteinase-1 (MMP1). Furthermore, we found inhibition of LPCAT1 ameliorates intestinal barrier dysfunction, diarrhea symptoms and colonic inflammation in LCA-treated mice and those with experimental colitis, highlighting LPCAT1 as a potential therapeutic target for gastrointestinal diseases characterized by intestinal barrier dysfunction and colonic inflammation. Additionally, our findings showed a positive correlation between LPCAT1 and biomarkers of intestinal barrier dysfunction and colonic inflammation in patients with ulcerative colitis. Collectively, these findings reveal the essential role of gut microbiota-derived LCA in the development of intestinal barrier dysfunction by activating LPCAT1, which worsens diarrhea symptoms in IBS and colonic inflammation in IBD. Inhibiting LPCAT1, therefore, presents a promising therapeutic strategy for both IBS-D and ulcerative colitis. This study provides insights into the molecular mechanisms involving LCA and an associated gut dysbiosis pattern in the pathogenesis of IBS-D, while also identifying new therapeutic targets aimed at maintaining intestinal homeostasis in gastrointestinal diseases, particularly in reducing IBD risks among the IBS-D population.

## Introduction

Irritable bowel syndrome (IBS) and inflammatory bowel disease (IBD) are two distinct gastrointestinal disorders, yet emerging evidence indicates a potential link between them. IBS is considered a functional disorder with no structural abnormalities, while IBD is characterized by chronic inflammation of the gastrointestinal tract(*1*). Epidemiological studies have shown a higher incidence of IBD in individuals previously diagnosed with IBS compared to the general population. For instance, a large population-based study revealed that patients with IBS had an increased risk of subsequently developing both Crohn’s disease and ulcerative colitis (UC), the two main subtypes of IBD(*2*), suggesting a possible association between IBS and the onset of IBD.

The intestinal barrier is composed of a single layer of epithelial cells lining the inner surface of the intestine, forming a physical and biochemical barrier between the gut lumen and the underlying tissue. It consists of several components, including tight junction proteins, mucus layers, antimicrobial peptides, and gut-associated lymphoid tissue(*3*). Disruption of intestinal barrier function has been implicated in various gastrointestinal disorders, including irritable bowel syndrome (IBS), inflammatory bowel disease (IBD) and colorectal cancer(*4*). Intestinal barrier dysfunction is commonly observed in both IBS and IBD and contributes to the progression of these conditions(*5*). Understanding the pathogenesis of intestinal barrier dysfunction is crucial for elucidating the increased incidence of IBD in IBS patients and for developing personalized prevention and treatment strategies for these complex gastrointestinal diseases.

Gut dysbiosis is an important pathogenic factor in IBS and IBD. Our previous studies showed a significant elevation of *clostridia* bacteria in IBS patients with excessive excretion of total bile acids (BAs), which contributes to diarrhea symptoms in IBS-D patients(*6*). However, it remains unclear whether microbial metabolism of BA is involved in the development of IBS-D. Secondary bile acids (SBAs) are produced by the gut microbiota through the metabolism of primary bile acids, such as cholic acid and chenodeoxycholic acid(*7*). Notably, SBAs, including deoxycholic acid and lithocholic acid, play a significant role in modulating intestinal barrier function. For instance, deoxycholic acid has been shown to disrupt tight junctions and increase intestinal permeability, leading to enhanced translocation of antigens and pathogens(*8*). Lithocholic acid (LCA) has been found to reduce the thickness of the mucus layer, compromising its protective function and allowing for increased contact between luminal contents and the epithelium(*9*). Therefore, alterations in SBAs due to gut dysbiosis may contribute to the pathogenesis of IBS and IBD by affecting intestinal barrier function.

In this study, we identified the biomarkers of intestinal barrier function in patients with IBS-D and characterized the association between intestinal barrier dysfunction and bile acids. We revealed LCA, a microbiota-derived bile acid, is positively correlated with intestinal barrier dysfunction in patients with IBS-D. Our experiments demonstrated that LCA disrupts intestinal barrier function in normal mice and aggravates intestinal inflammation in a dextran sulfate sodium (DSS)-induced colitis model. Through lipidomics analysis, mouse studies and *in vitro* studies, we revealed that LCA significantly alters phospholipid metabolism and directly activates LPCAT1 in the colonic tissues of mice. Accordingly, we investigated the regulatory role of LPCAT1 in intestinal barrier function and found that its overexpression induces intestinal barrier dysfunction and promotes pro-inflammatory responses via MMP1. In contrast, inhibition of LPCAT1 improves intestinal barrier dysfunction, diarrhea symptoms, and colonic inflammation in mouse models with impaired intestinal barrier function induced by LCA and DSS, respectively. Finally, we validated the clinical relevance of LPCAT1 in intestinal barrier dysfunction in IBD by demonstrating a positive correlation between LPCAT1 and biomarkers of intestinal barrier dysfunction and colonic inflammation in patients with UC.

## Results

### LCA is positively correlated with intestinal barrier dysfunction in patients with IBS-D

We first determined the widely recognized biomarkers of intestinal barrier function including lipopolysaccharide-binding protein (LBP) and intestinal fatty acid binding protein (I-FABP)(*10*) in the serum samples of IBS-D patients and healthy controls (HC). As shown in (p<0.05 in all cases, Figure.S1A-B), both serum LBP and I-FABP levels were significantly increased in IBS-D patients, revealing that intestinal barrier dysfunction was present in approximately 31% of IBS-D patients using a 90% cut-off value of I-FABP from HC subjects. Furthermore, we showed that I-FABP level was positively correlated with the severity of gastrointestinal symptoms, including diarrhea and abdominal pain in patients with IBS-D (r=0.19 and 0.21, p<0.05 in all cases, Figure.S1C), suggesting that intestinal barrier dysfunction is prevalent in IBS-D and contributes to the development of gastrointestinal (GI) symptoms.

Our previous study indicated that gut dysbiosis is prevalent in IBS-D patients, with total BA overexcretion driven by a *Clostridia*-rich microbiota contributing to diarrhea symptoms in IBS-D(*11*). Consequently, we conducted a correlation analysis to address the clinical relevance between BA profiles and intestinal barrier dysfunction in patients with IBS-D. We then determined the BA profiles in serum samples from IBS-D patients and HC, regardless of excess BA excretion, which differs from our previous study(*6*). Through correlation analysis between serum BA profiles with the I-FABP index, we demonstrated gut microbiota-derived LCA is positively correlated with biomarkers of intestinal barrier dysfunction (I-FABP and LBP indexes), showing the highest correlation coefficient among all BAs (Figure.1A). Both primary and secondary BAs were significantly altered in the serum of IBS-D patients compared to HC, revealing that both synthetic and metabolic pathways of bile acids are significantly altered in IBS-D patients (Figure.1B). Among the altered BAs, we also observed that LCA levels were significantly increased in the serum of IBS-D patients (p<0.0001, Figure.1B). Specifically, LCA elevation was found in 61.4% IBS-D patients using a 90% cut-off value from HC subjects.

**Figure.1.**
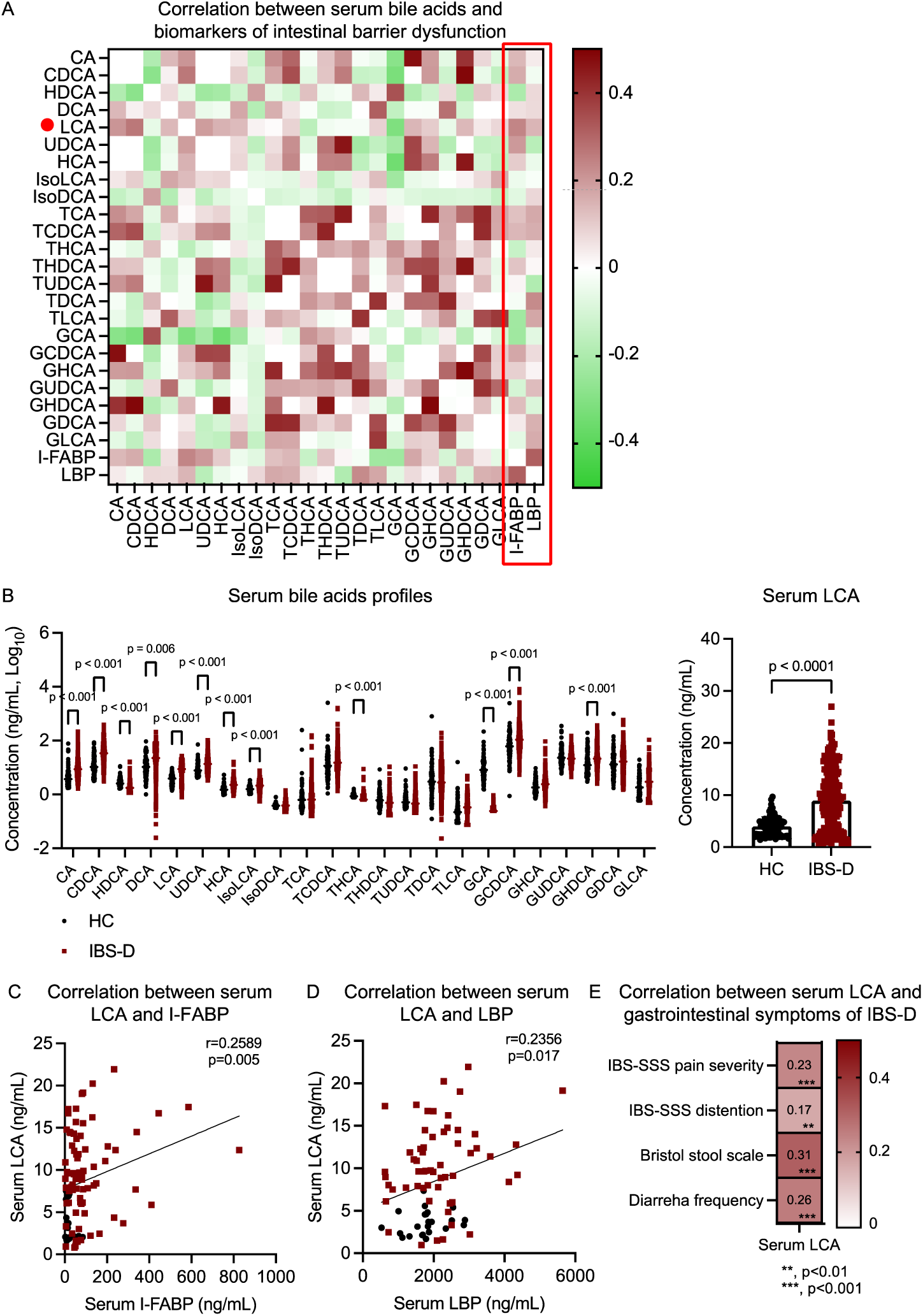
LCA is positively associated with intestinal barrier dysfunction and GI symptoms in patients with IBS-D. **(A)** Nonparametric spearman correlation analysis between serum bile acids profiles and biomarkers of intestinal barrier dysfunction (serum I-FABP and LBP) in IBS-D patients (one-tailed). **(B)** Serum bile acids profiles and detailed changes in serum LCA in IBS-D patients (n=248) and healthy controls (HC, n=76). Data were analyzed using a two-tailed t-test. **(C-D)** Nonparametric spearman correlation analysis between serum LCA and biomarkers of intestinal barrier dysfunction (serum I-FABP and LBP) in HC and IBS-D patients (one-tailed). **(E)** Nonparametric spearman correlation analysis between serum LCA and the severity of GI symptoms in IBS-D patients (one-tailed).

We then performed correlation analysis to explore the detailed relationship between clinical indexes of GI symptoms in IBS-D patients, serum LCA levels, and intestinal barrier function-related biomarkers. As shown in Figure.1C-D, (r=0.2589 and 0.2356, p<0.05 in all cases), both LCA and intestinal barrier function biomarkers, including I-FABP and LBP indexes, were positively associated with the severity of GI symptoms of IBS-D patients including abdominal pain score, abdominal distention, Bristol stool scale, and defecation frequency (r=0.23, 0.17, 0.31 and 0.26, p<0.01 in all cases, Figure.1E). Our results suggested a positive correlation among gut dysbiosis-induced LCA elevation, intestinal barrier dysfunction, and GI symptoms in IBS-D patients, suggesting that LCA is linked to intestinal barrier dysfunction in IBS-D patients, thus contributing to the development of GI symptoms.

### LCA triggers intestinal barrier dysfunction in mice

We then investigated the effects of LCA within a pathophysiological dosage on intestinal barrier function in mice (Figure.2A). We showed both colonic LCA (1 µM) and serum LCA (200 nM) were significantly elevated in LCA-treated mice compared with control group (p<0.001, Figure 2B-C). Moreover, serum levels of I-FABP and FITC-dextran, biomarkers for intestinal barrier integrity, were significantly increased in mice receiving LCA treatment (p<0.01, Figure.2D-E). In line with this finding, we observed an increased colonic permeability in colonic tissues of LCA-treated mice measured by the Ussing chamber method (p<0.001, Figure.2F), revealing LCA disrupts intestinal barrier function in normal mice. Moreover, we showed an increased fecal water content and an accelerated defecation frequency in normal mice receiving LCA treatment (p<0.05, Figure.2G-H), suggesting that excessive LCA contributes to the diarrhea symptoms of IBS-D. No significant changes in body weight and colon length were found in the normal mice treated with LCA (n.s., Figure.2I-J). Histological analysis of colon tissues using H&E and Alcian blue staining revealed increased infiltration of inflammatory cells and disrupted goblet cell architecture in LCA-treated mice (p<0.01, Figure.2K). We further validated the detrimental effects of LCA on intestinal barrier function by measuring permeability in Caco-2 cells. As expected, LCA treatment at physiological dosages impaired intestinal barrier function, as indicated by increased permeability of Caco-2 cells in a dose-dependent manner (p<0.05, Figure.2L-M).

**Figure.2.**
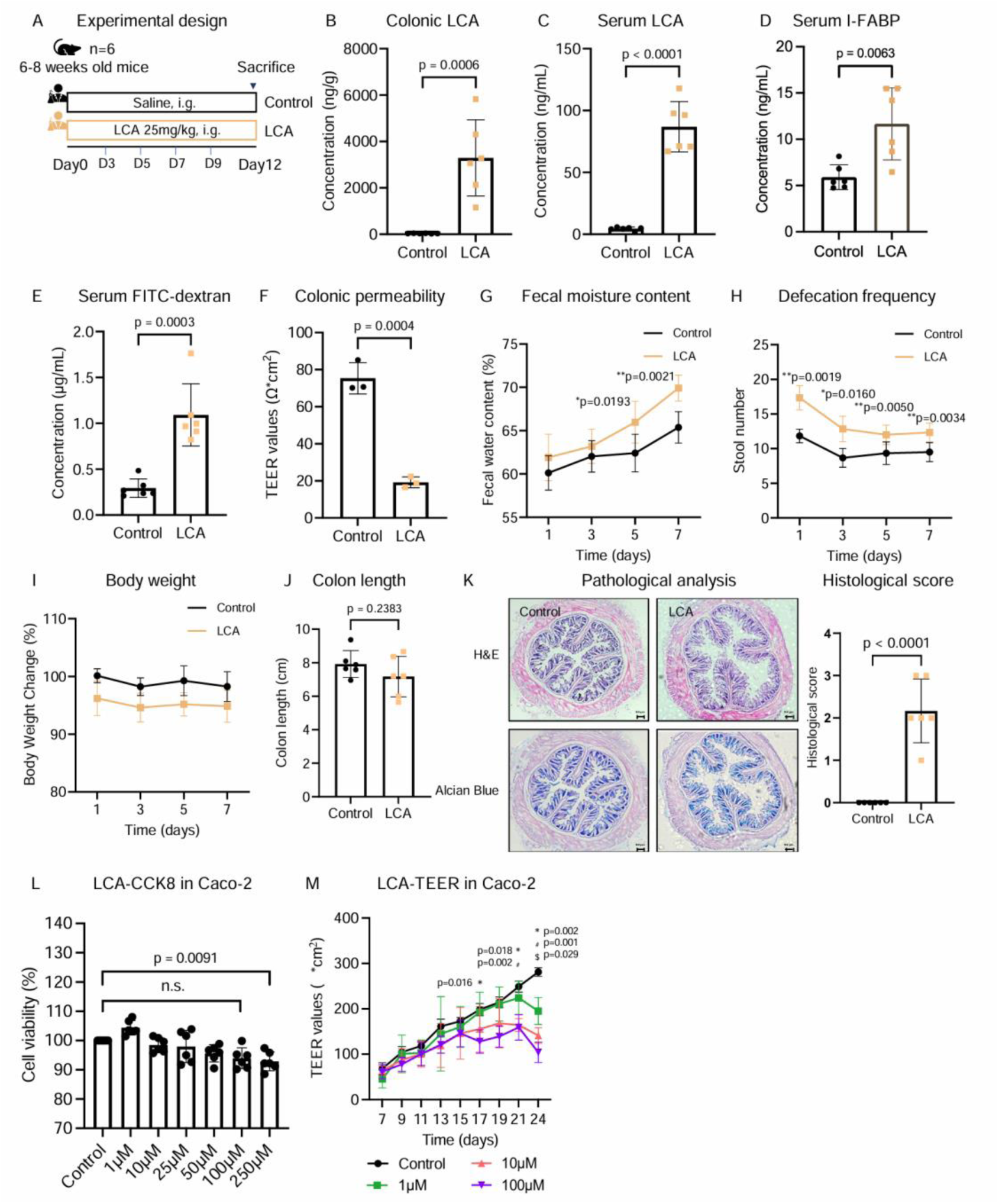
LCA triggers intestinal barrier dysfunction and diarrhea-like symptoms in normal mice and impairs intestinal barrier integrity in Caco-2 cells. **(A)** Schematic illustration of evaluation of LCA action on intestinal barrier function and GI motility in normal mice. **(B-C)** Colonic and serum levels of LCA in mice treated with vehicle control or LCA (n=6 per group). **(D-E)** Serum levels of I-FABP and FITC-Dextran in mice treated with vehicle control or LCA (n=6 per group). **(F)** Colonic barrier permeability in mice treated with vehicle control or LCA (n=3 per group). **(G-J)** Fecal water content, defecation frequency, body weight changes and colon length in mice treated with vehicle control or LCA (n=6 per group). **(K)** Pathological analysis of H&E and Alcian blue staining of colon tissues from mice treated with vehicle control or LCA (n=6 per group). **(L)** Cell viability of Caco-2 cells treated with various concentrations of LCA (n=3 per group). **(M)** Cell permeability of Caco-2 cells monolayer model treated with various concentrations of LCA (n=3 per group). Data were analyzed using either two-tailed t-test or two-tailed two-way ANOVA.

We then utilized a mouse model of colonic inflammation induced by DSS further investigate the effects of LCA on the exacerbation of intestinal barrier dysfunction and colonic inflammation (Figure.3A). In alignment with previous findings in normal mice, LCA treatment resulted in significantly elevated levels of LCA in both colonic tissues and serum (p<0.001, Figure 3B-C), in accompanied with a significant worsening of intestinal barrier dysfunction (p<0.05 in all cases, Figure.3D-F), GI symptoms (p<0.05 in all cases, Figure.3G-I) and colonic inflammation (p<0.05, Figure.3J) in DSS-treated mice, as indicated by biomarkers of intestinal barrier dysfunction (I-FABP and FITC-dextran), colonic permeability, fecal water content, colon length changes, disease activity index (DAI) and H&E and Alcian blue staining. Taken together, these results suggest that LCA contributes to intestinal barrier dysfunction and aggravates colonic inflammation in DSS-treated mice.

**Figure.3.**
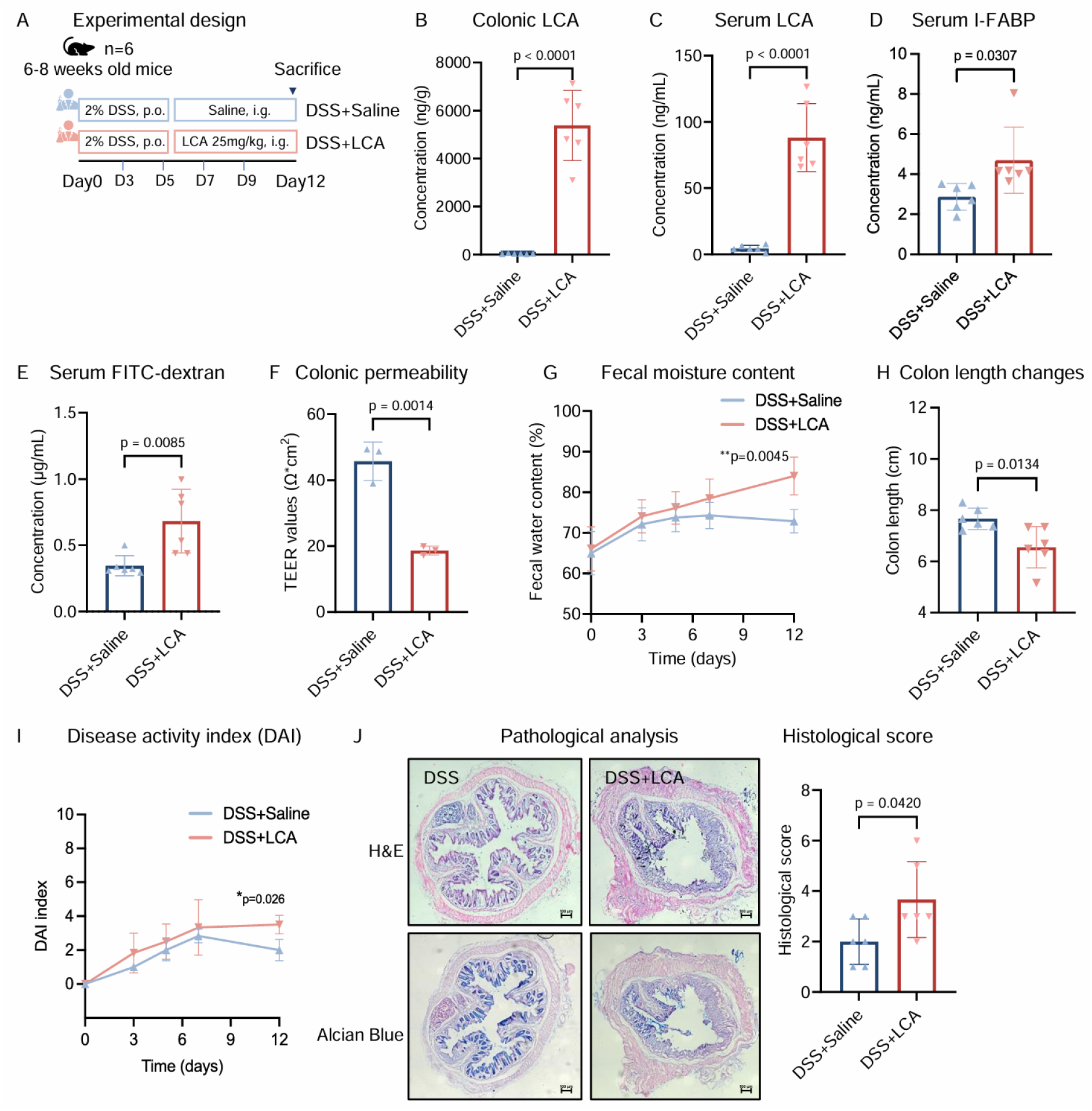
LCA aggravates intestinal barrier dysfunction and colonic inflammation in a mouse model of colitis. **(A)** Schematic illustration of evaluation of LCA action on intestinal barrier function and disease severity in a mouse model of colitis induced by DSS. **(B-C)** Colonic and serum levels of LCA in DSS-treated mice co-treated with vehicle control or LCA (n=6 per group). **(D-E)** Serum levels of I-FABP and FITC-Dextran in DSS-treated mice co-treated with vehicle control or LCA (n=6 per group). **(F)** Colonic barrier permeability in mice treated with vehicle control or LCA (n=3 per group). **(G-I)** Fecal water content, colon length and DAI score in DSS-treated mice co-treated with vehicle control or LCA (n=6 per group). **(J)** Pathological analysis of H&E and Alcian blue staining of colon tissues from DSS-treated mice co-treated with vehicle control or LCA (n=6 per group). Data were analyzed using either two-tailed t-test or two-tailed two-way ANOVA.

### LCA alters phospholipid profiles and upregulates LPCAT1 expression in mice

Phospholipids are important components of the intestinal mucosal barrier(*12, 13*). In this study, we analyzed the lipidome of colonic tissues from LCA-treated mice using lipidomics to investigate the underlying mechanisms by which LCA impairs intestinal barrier function. We showed lipidome signatures were significantly altered in the colonic tissues of LCA-treated mice (Figure.S2A-F). Moreover, we revealed the levels of phosphatidylcholine (PC) were significantly upregulated in the colonic tissues of LCA-treated mice under both normal and inflammatory conditions (p<0.05, Figure.4A-B), suggesting LCA action on intestinal barrier function is associated with the changes in PC. As LPCAT is responsible for the transformation of lysophospholipids to phospholipids in the remodeling pathway of PC biosynthesis(*14*), we determined the mRNA expression changes of LPCAT (LPCAT1-4) in the colonic tissues of LCA-treated mice and we showed that the LPCAT1 expression was exclusively upregulated by LCA (p<0.01, Figure.4C-D).

**Figure.4.**
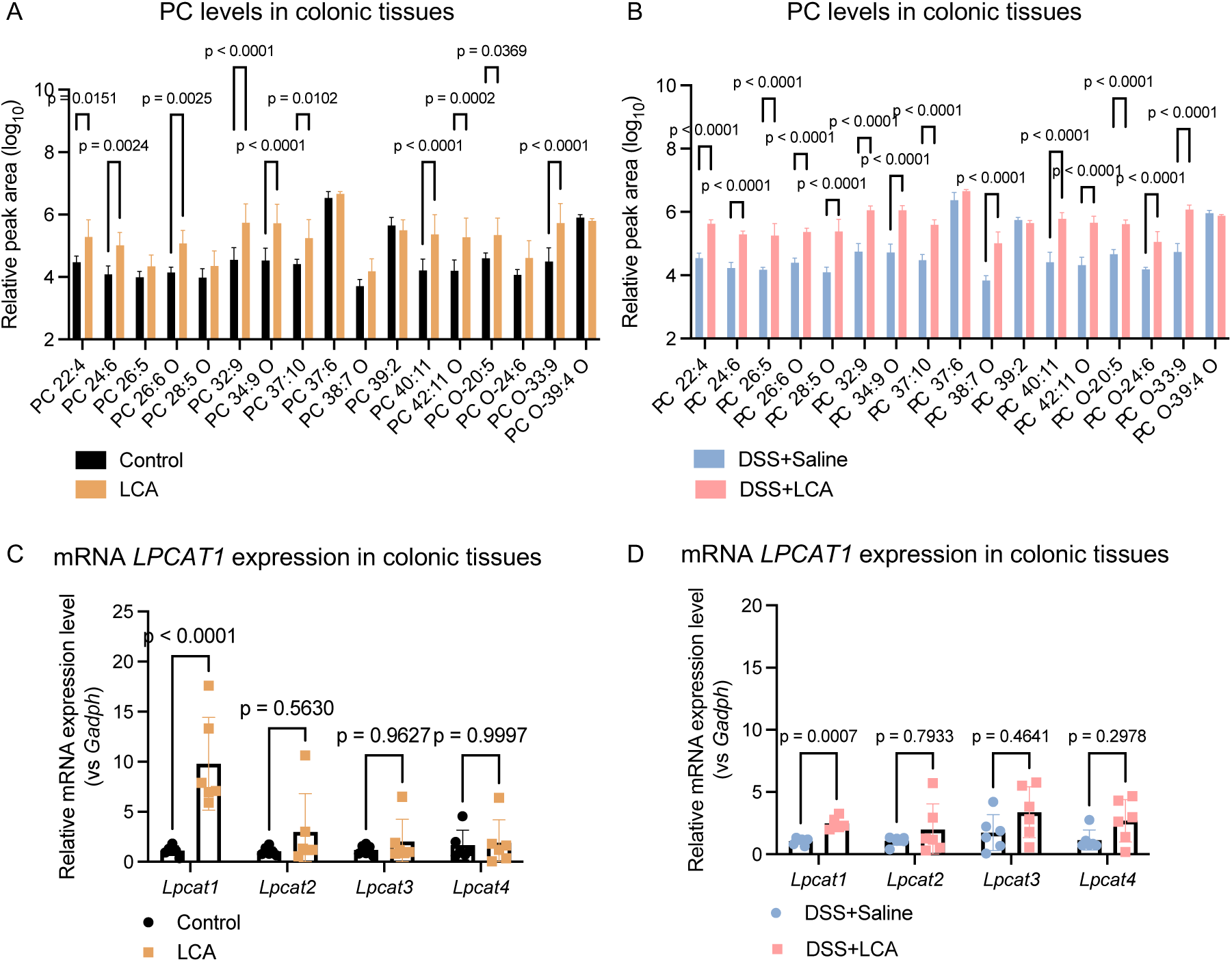
LCA upregulates PC levels and LPCAT1 expression in normal and DSS-treated mice. **(A-B)** Colonic PC levels in normal and DSS-treated mice co-treated with vehicle control or LCA (n=6 per group). **(C-D)** Colonic mRNA expression of *Lpcat1, Lpcat2, Lpcat3 and Lpcat4* in normal and DSS-treated mice co-treated with vehicle control or LCA (n=6 per group). Data were analyzed using a two-tailed two-way ANOVA.

In line with *in vivo* lipidomics findings, our *in vitro* observations showed that treatment of LCA altered phospholipid profiles including PC levels and selectively upregulated LPCAT1 expression in LCA-treated Caco-2 cells (Figure.S3A-F). Collectively, these results suggest that LPCAT1 activation is a critical mechanism involved in gut dysbiosis/LCA-induced intestinal barrier dysfunction in IBS-D.

### LCA is an endogenous activator of LPCAT1

To investigate the underlying mechanisms by which LCA activates LPCAT1, we assessed the stability of LPCAT1 in the presence of LCA in Caco-2 cells using thermal shift assays and cycloheximide (CHX) chase assays. In the temperature-dependent stability test, our results demonstrated that LPCAT1 exhibited increased thermal stability when incubated with LCA, indicating that LCA enhances the structural integrity of LPCAT1 (p<0.05, Figure.5A). In the CHX chase assay, LPCAT1 levels in LCA-treated Caco-2 cells were significantly higher than in untreated controls, indicating that LCA prevents LPCAT1 degradation (p<0.05, Figure.5B). We then employed microscale thermophoresis (MST) to quantify the binding affinity between LPCAT1 and LCA. The MST analysis revealed a strong interaction between LPCAT1 and LCA with a dissociation constant (K_d_) of 4.829 µM. In contrast, deoxycholic acid (DCA) exhibited a weak affinity (K_d_=259.7 µM), while chenodeoxycholic acid (CDCA) showed no affinity to LPCAT1 (Figure.5C). These findings indicated that microbiota-derived LCA acts as an endogenous agonist of LPCAT1, facilitating its activation exclusively.

**Figure.5.**
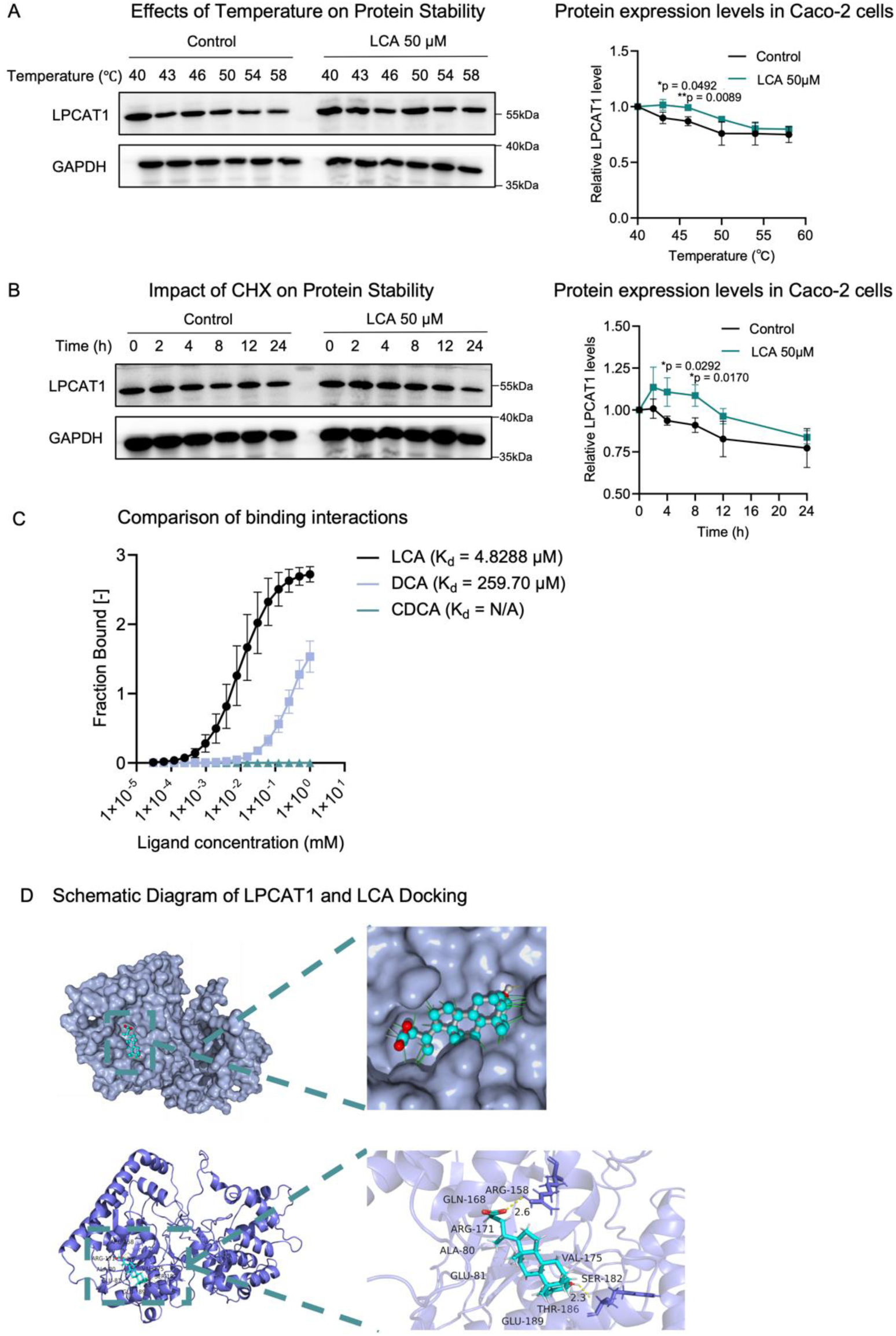
LCA is an endogenous activator of LPCAT1. **(A)** Western blot analysis and semi-quantification of LPCAT1 in Caco-2 cells treated with vehicle control or LCA at indicated temperature (n=3 per group). **(B)** Western blot analysis and semi-quantification of LPCAT1 in CHX-treated Caco-2 cells co-treated with vehicle control or LCA at indicated time points (n=3 per group). **(C)** The binding affinity of LCA, DCA and CDCA for LPCAT1 at indicated concentration. **(D)** Illustration of the molecular docking interactions between LPCAT1 and LCA. Data were analyzed using a two-tailed two-way ANOVA

Accordingly, we performed molecular docking studies to elucidate the interactions between LCA and LPCAT1. The docking analysis predicted a binding conformation of LCA within the binding sites of LPCAT1, involving key amino acid residues such as Arg158, Gln168, and Tyr186 (Figure.5D). These interactions likely contribute to the enhanced stability and activation of LPCAT1 upon LCA binding, providing a mechanistic understanding of how LCA activates LPCAT1. These results demonstrated that LCA acts as an endogenous agonist of LPCAT1, enhancing its stability and function. The high-affinity binding and structural insights highlight the significance of LCA-LPCAT1 interactions in the development of intestinal barrier dysfunction and colonic inflammation.

### LCA induces pro-inflammatory protease MMP1 secretion via LPCAT1

As the regulatory role of LPCAT1 in intestinal barrier function has not been studied before, we used a transcriptome approach to determine the changes in mRNA expression in Caco-2 cells overexpressing LPCAT1 under both normal and pro-inflammatory conditions (Figure.S4A-B). We demonstrated that MMP1, a matrix metalloproteinase protein that disrupts the intestinal epithelial barrier by processing pro-inflammatory cytokine(*15–18*), was significantly increased among the altered genes in both wild-type and LPCAT1-overexpressing Caco-2 cells under normal and inflammatory conditions (p<0.05, Figure.S4C-D). Additionally, we showed that LCA treatment upregulated the protein expression of LPCAT1 and MMP1 in Caco-2 cells (p<0.01, Figure.S4E), demonstrating that LCA induces MMP1 secretion and LPCAT1 upregulation. In line with our *in vitro* studies, we also observed significant increases in the protein expression of both LPCAT1 and MMP1 in the colonic tissues of mouse models with intestinal barrier dysfunction, including those treated with LCA treatment and those with DSS-induced experimental colitis (p<0.05 in all cases, Figure.6A-C).

Moreover, we showed that the expression of LPCAT1 and MMP1 is significantly reduced in germ-free mice, accompanied by depleted levels of secondary bile acids compared to specific pathogen-free (SPF) mice, indicating that both LPCAT1 and MMP1 are predominantly regulated by gut microbiota-derived metabolites (p<0.05, Figure.6D-E). Given that LCA upregulates the expression levels of both LPCAT1 and MMP1, a microbiota-dependent LCA-LPCAT1-MMP1 axis is implicated in the pathogenesis of intestinal barrier dysfunction in IBS-D.

**Figure.6.**
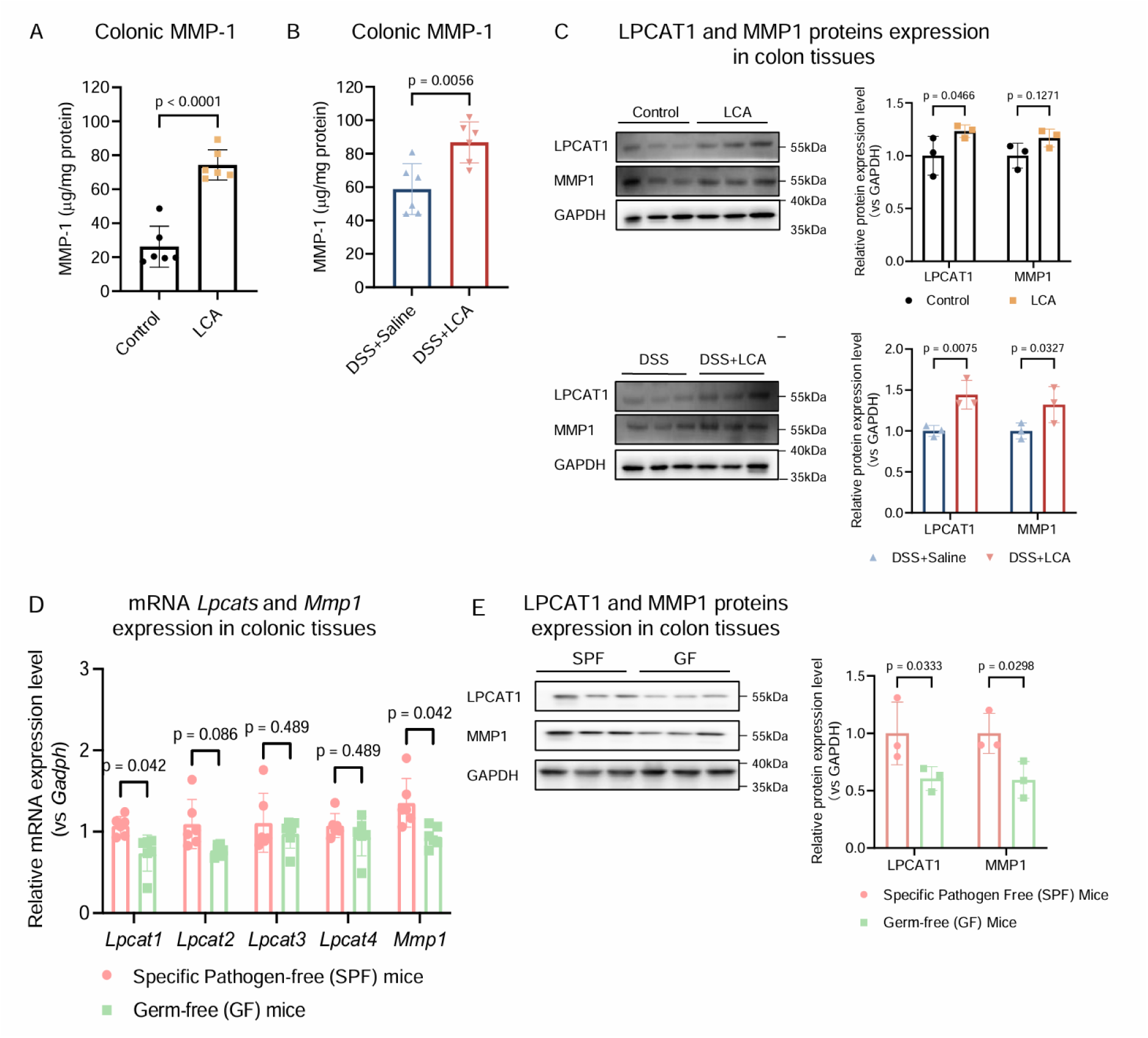
LPCAT1-MMP1 is suppressed by gut microbiota depletion and upregulated by gut microbiota-derived LCA. **(A-B)** Colonic MMP-1 levels in normal and DSS-treated mice co-treated with vehicle control or LCA (n=6 per group). **(C)** Western blot analysis and semi-quantification of LPCAT1 and MMP1 in colonic tissues from normal mice and DSS-treated mice co-treated with vehicle control or LCA (n=6 per group). **(D)** mRNA expression levels of *Lpcat1, Lpcat2, Lpcat3, Lpcat4* and *Mmp1* in colonic tissues from SPF mice and GF mice. **(E)** Western blot analysis and semi-quantification of LPCAT1 and MMP1 in colonic tissues from SPF mice and GF mice (n=3 per group). Data were analyzed using a two-tailed two-way ANOVA.

### LPCAT1-MMP1 impairs intestinal barrier function and exacerbates pro-inflammatory responses in a positive feedback loop

To study the role of LPCAT1 in intestinal barrier dysfunction, we established LPCAT1^flox/-^ (CKI) mice and conditional knock-in LPCAT1^flox/flox^ (CRE) mice to examine the pathological changes under experimental colitis conditions (Figure.7A). Notably, we observed that body weight loss, DAI score elevation, and colon length shortening were significantly aggravated in LPCAT1^flox/flox^ (CRE) mice under challenge of DSS-induced colitis (p<0.05 in all cases, Figure.7B-D). In parallel, we found that colonic injuries and intestinal barrier integrity were significantly worsened in the colonic tissues of LPCAT1^flox/flox^ (CRE) mice, as revealed by H&E staining and IF staining of ZO-1 and occludin (p<0.05, Figure.7E-F). Our findings were further supported by the determination of intestinal barrier function biomarkers, including serum FD4 and colonic ZO-1, occludin and claudin-2, as well as pro-inflammatory factors including colonic MMP-1, TNF-α and IL-6 (p<0.05 in all cases, Figure.7G-K). Collectively, all of these biological indexes demonstrated that intestinal barrier dysfunction is aggravated in LPCAT1^flox/flox^ (CRE) mice, indicating that LPCAT1 activation by LCA disrupts intestinal barrier function and aggravates colonic inflammatory responses in experimental colitis.

**Figure.7.**
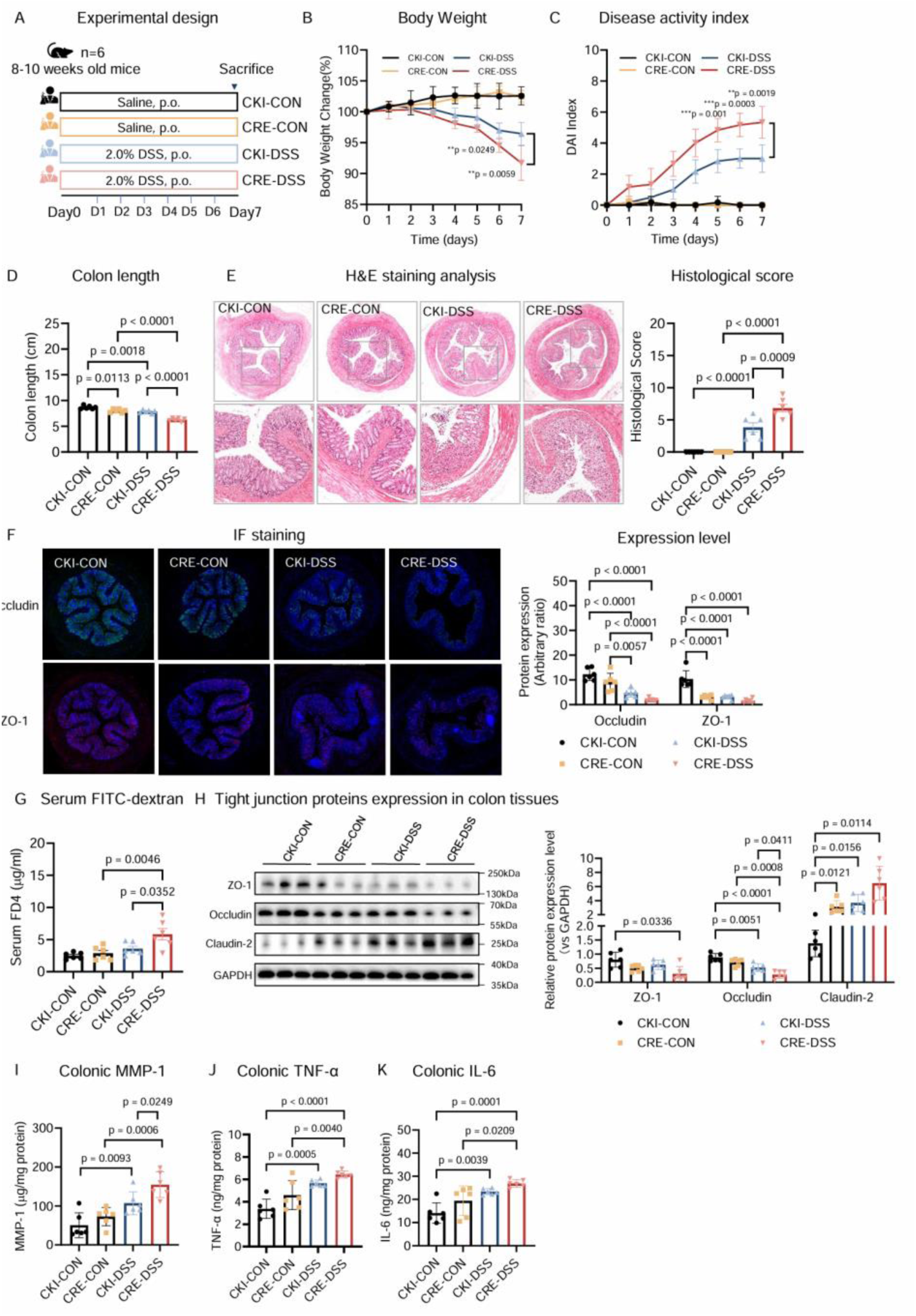
LPCAT1 overexpression impairs intestinal barrier function and exacerbates colonic inflammation in mice. **(A)** Schematic illustration of evaluation of LPCAT1 overexpression on intestinal barrier function, GI motility and colonic inflammation in Rosa26-LSL-LPCAT1^flox/-^ mice (CKI mice) and villin-Cre LSL-LPCAT1^flox/flox^ mice (CRE mice)**. (B-D)** Body weight, DAI score and colon length in CKI and CRE mice treated with water or DSS (n=6 per group). **(E-F)** Pathological analysis including H&E staining and IF staining of Occludin and ZO-1 in colonic tissues of CKI and CRE mice treated with water or DSS (n=6 per group). **(G)** Serum FITC-Dextran in CKI and CRE mice treated with water or DSS (n=6 per group). Data were analyzed using a two-tailed one-way ANOVA. **(H)** Western blot analysis and semi-quantification of ZO-1, Occludin and Claudin 2 in colonic tissues of CKI and CRE mice treated with water or DSS (n=3 per group). **(I-K)** Colonic MMP-1, TNF-α and IL-6 in CKI and CRE mice treated with water or DSS (n=6 per group). Data were analyzed using a two-tailed one-way ANOVA.

In line with our *in vivo* observations, we demonstrated that LPCAT1 overexpression leads to impaired intestinal permeability, increased markers of intestinal barrier dysfunction and pro-inflammatory factors, and a reduced wound healing rate (p<0.05 in all cases, Figure.S5A-E) in Caco-2 cells under both normal and inflammatory stimulation. We also validated the role of MMP1 in maintaining intestinal barrier integrity, showing that MMP1 recombinant protein (MMP1 RP) disrupts intestinal barrier integrity, as evidenced by impaired intestinal permeability, elevated markers of intestinal barrier dysfunction, and a decreased wound healing rate (p<0.05 in all cases, Figure.S6A-E). Collectively, these findings indicate that LPCAT1 activation leads to intestinal barrier dysfunction and aggravates pro-inflammatory responses. Moreover, MMP1 secretion induced by LPCAT1 creates a positive feedback loop, whereby intensifying the pro-inflammatory response in the gut and impairing intestinal barrier function.

### Inhibiting LPCAT1-MMP1 restores intestinal barrier dysfunction and alleviates colonic inflammation

We then investigated whether inhibition of LPCAT1 and MMP1 by gene knock-out approaches restored intestinal barrier permeability and pro-inflammatory responses in Caco-2 cells. We found that both MMP1 and LPCAT1 gene knock-down improved impaired intestinal permeability and pro-inflammatory responses in both wild-type and LPCAT1 overexpressed Caco-2 cells under TNF-α treatment (p<0.05 in all cases, Figure.S7A-C). Additionally, we observed mRNA expression levels of LPCAT1 were downregulated, and intestinal barrier dysfunction and pro-inflammatory responses were suppressed following MMP1 gene knock-down in LPCAT1 overexpressed Caco-2 cells under TNF-α treatment (Figure.S7D-E). In line with these findings, we showed impaired intestinal barrier permeability, LPCAT1-MMP1 upregulation and a decreased wound healing rate were significantly reversed by treatment of TS1-01 (a selective LPCAT1 inhibitor) in TNF-α-treated Caco-2 cells (p<0.05 in all cases, Figure.S8A-E). These results showed that inhibition of the LPCAT1-MMP1 axis using pharmacological inhibition and gene knock-out approaches restores intestinal barrier permeability and mitigates pro-inflammatory responses

We further investigated whether inhibition of LPCAT1 by pharmacological inhibition restored intestinal barrier dysfunction and colonic inflammation in DSS-treated mice (Figure.8A) and LCA-treated mice (Figure.9A). In line with *in vitro* observations, we demonstrated colonic injuries and pro-inflammatory responses in DSS-induced colitic mice were significantly ameliorated by the treatment of TS1-01 (p<0.05 in all cases, Figure.8B-E). Moreover, TS1-01 treatment significantly improved biomarkers of intestinal barrier dysfunction and downregulated colonic levels of MMP-1 and LPCAT1 in DSS-induced colitic mice (p<0.05 in all cases, Figure.8F-K). In alignment with our findings in the colitis model, we showed that LCA-induced intestinal barrier dysfunction and diarrhea-like symptoms were also significantly improved by the treatment of TS1-01 (p<0.05 in all cases, Figure.9B-L). These results support the notion that LPCAT1-MMP1 is a potential target for treating intestinal barrier dysfunction, thereby alleviating diarrhea symptoms in IBS-D.

**Figure.8.**
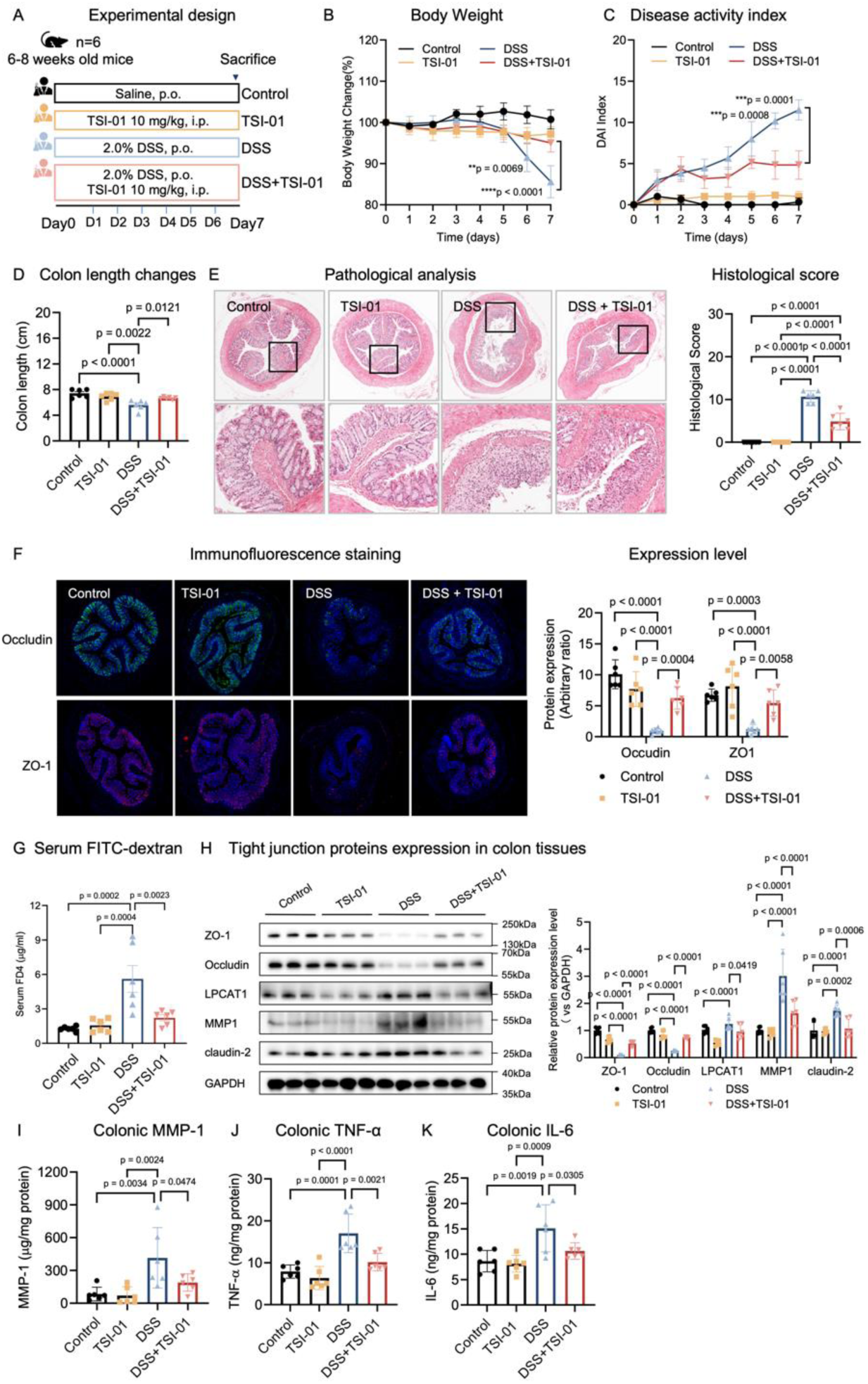
Inhibiting LPCAT1-MMP1 by an LPCAT1 inhibitor TSI-01 alleviates intestinal barrier dysfunction and colonic inflammation in DSS-treated mice. **(A)** Schematic illustration of evaluation of TSI-01 action on intestinal barrier function and disease severity in a mouse model of colitis induced by DSS. **(B-D)** Body weight, DAI score and colon length in DSS-treated mice co-treated with vehicle control or TSI-01 (n=6 per group). **(E-F)** Pathological analysis including H&E staining and IF staining of Occludin and ZO-1 in colonic tissues from DSS-treated mice co-treated with vehicle control or TSI-01 (n=6 per group). **(G)** Serum FITC-Dextran in DSS-treated mice co-treated with vehicle control or TSI-01 (n=6 per group). **(H)** Western blot analysis and semi-quantification of LPCAT1, MMP1, ZO-1, Occludin and Claudin 2 in colonic tissues from DSS-treated mice co-treated with vehicle control or TSI-01 (n=6 per group). **(I-K)** Colonic MMP-1, TNF-α and IL-6 in DSS-treated mice co-treated with vehicle control or TSI-01 (n=6 per group). Data were analyzed using a two-tailed one-way ANOVA.

**Figure.9.**
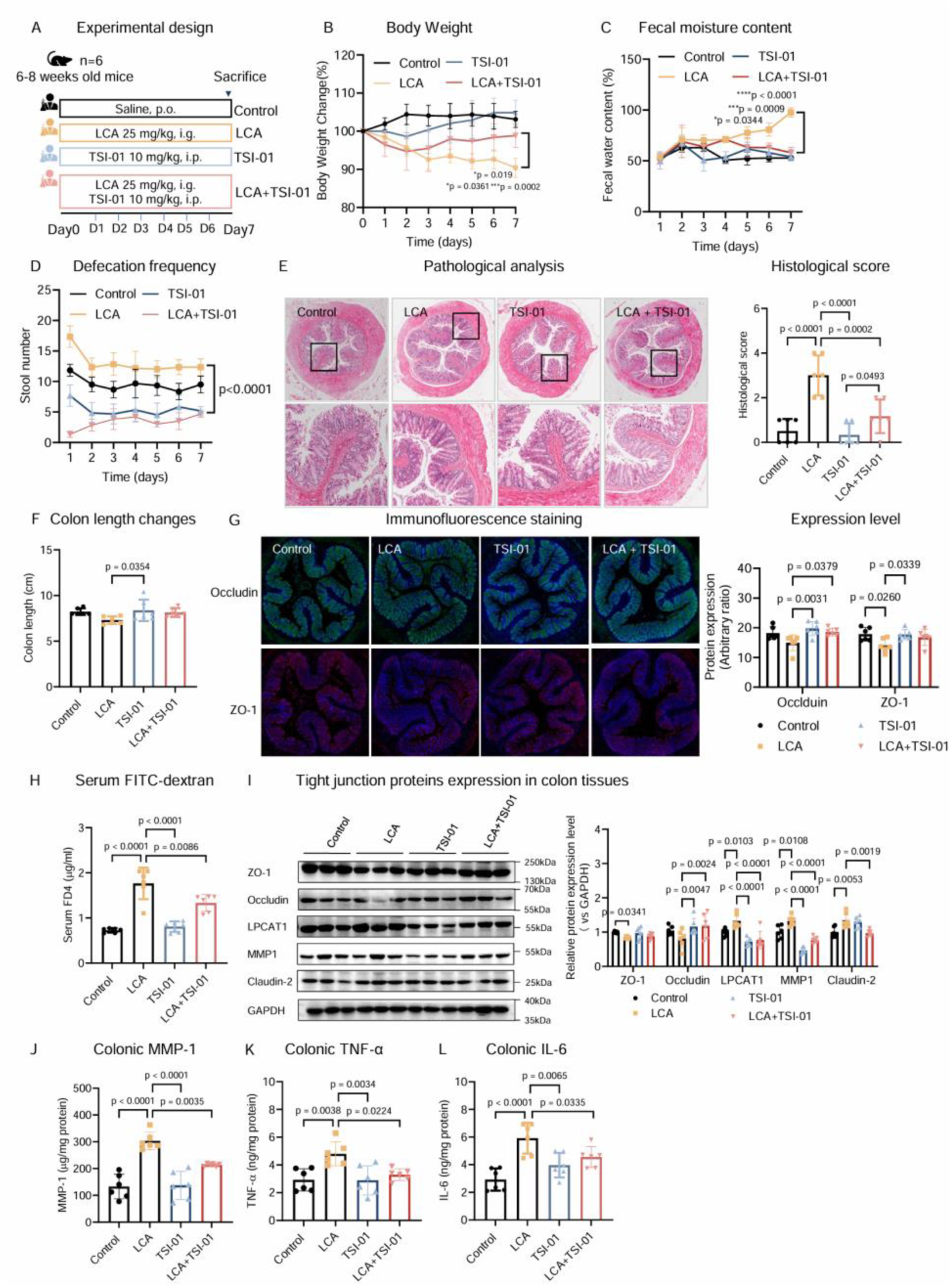
Inhibiting LPCAT1-MMP1 by an LPCAT1 inhibitor TSI-01 alleviates intestinal barrier dysfunction and colonic inflammation in LCA-treated mice. **(A)** Schematic illustration of evaluation of TSI-01 action on intestinal barrier function and disease severity in LCA-treated mice. **(B-D)** Body weight changes, fecal water content and defecation frequency in LCA-treated mice co-treated with vehicle control or TSI-01 (n=6 per group). **(E, G)** Pathological analysis including H&E staining and IF staining of Occludin and ZO-1 in colonic tissues from LCA-treated mice co-treated with vehicle control or TSI-01 (n=6 per group). **(F)** Colon length in LCA-treated mice co-treated with vehicle control or TSI-01 (n=6 per group). **(H)** Serum FITC-Dextran in LCA-treated mice co-treated with vehicle control or TSI-01 (n=6 per group). **(I)** Western blot analysis and semi-quantification of LPCAT1, MMP1, ZO-1, Occludin and Claudin 2 in colonic tissues from LCA-treated mice co-treated with vehicle control or TSI-01 (n=6 per group). **(J-L)** Colonic MMP-1, TNF-α and IL-6 in LCA-treated mice co-treated with vehicle control or TSI-01 (n=6 per group). Data were analyzed using a two-tailed one-way ANOVA.

### LPCAT1 is positively correlated with intestinal barrier dysfunction in patients with UC

Due to the fact that colonic tissue biospecimens are not available to collect from patients with IBS-D and healthy controls in this study, we investigate the mRNA and protein expression of LPCAT1 and biomarkers of intestinal barrier dysfunction using colonic tissue biospecimens from patients with UC to further address the clinical relevance of LPCAT1 in intestinal barrier dysfunction. We determined LPCAT1 expression in the inflamed colon tissues of patients with UC in Gene Expression Omnibus (GEO) datasets (GSE6731(*19*), GSE9452(*20*), GSE22619(*21*) and GSE38713(*22*)) and validated the results in patients with UC. Notably, we found that both mRNA expression and immunostaining of LPCAT1 were significantly increased in colonic tissues from different cohorts of patients with UC (p<0.01, Figure.S9A). We also observed a positive correlation between LPCAT1 and biomarkers of intestinal barrier dysfunction Claudin 2 and Occludin (p<0.01 in all cases, Figure.S9B). We also showed that the LPCAT1 expression was significantly increased in colonic tissues from UC patients with both inactive and active stages compared with marginal normal tissue excised from patients with colon tumors and positively correlated with colonoscopy score based on immunohistochemistry analysis (r=0.4668, p<0.01, Figure.S9C-D). Moreover, we showed LPCAT1 expression was positively correlated with Claudin 2, a biomarker of intestinal barrier dysfunction, in colonic tissues from patients with UC (r=0.5417, p<0.05, Figure.S9E). In summary, LPCAT1 is significantly increased in UC patients and positively correlated with intestinal barrier dysfunction and disease severity.

## Discussion

Intestinal barrier dysfunction is a significant characteristic in GI diseases, including IBS and IBD, as it contributes to system inflammation and aggravates GI symptoms such as altered bowel movements and abdominal pain(*23*). A systematic review indicated that intestinal barrier dysfunction is prevalent in both adult and pediatric IBS studies, particularly in the IBS-D subtype(*24*). Moreover, a positive correlation between the loss of intestinal barrier function and the severity of GI symptoms has been observed in these IBS studies, aligning with our current findings in IBS-D patients. As a common feature in both IBS and IBD, intestinal barrier dysfunction facilitates the translocation of bacteria, toxins, and antigens into the mucosa and systemic circulation. Therefore, maintaining intestinal barrier function holds significant potential as a novel management strategy for IBS and IBD.

In this study, we demonstrated that elevated levels of gut-microbial LCA induced by gut dysbiosis disrupt intestinal barrier function via LPCAT1, leading to diarrhea symptoms in IBS-D and colonic inflammation in IBD. Inhibition of LPCAT1 effectively ameliorated intestinal barrier dysfunction, alleviated diarrhea-like symptoms and reduced colonic inflammation in both LCA-treated and DSS-treated mice, suggesting it is a promising therapeutic target for IBS and IBD. Furthermore, our clinical data showed a positive association between LCA, LPCAT1, intestinal barrier dysfunction and GI symptoms in patients with IBS and IBD. This study identifies LCA as a gut microbiota-derived pathogenic factor for IBS-D and IBD, and highlights LPCAT1 as a therapeutic target to protect against diarrhea-like symptoms and colonic inflammation in IBS-D and IBD by maintaining intestinal barrier function.

Gut dysbiosis, as widely found in both IBS and IBD together with intestinal barrier dysfunction, plays a key role in the pathogenesis of these conditions. The altered gut microbiota can directly damage the barrier or manipulate the production of host and gut-microbial metabolites and proteins which are critical for the biological functions of intestinal epithelial cells. For IBS-D, gut dysbiosis has been implicated in the GI symptoms by our previous studies(*25, 26*), and the role of intestinal barrier dysfunction induced by gut dysbiosis in IBS-D is well answered in this study. For IBD, gut dysbiosis not only contributes to intestinal barrier dysfunction but also inappropriate immune activation in the context of a “leaky gut”(*27*). Therefore, targeting the improvement of gut microbiota and barrier function by drugs, probiotics, prebiotics, and dietary modifications can effectively relieve GI symptoms and maintain immune homeostasis.

LCA, a secondary bile acid produced by the microbial metabolism of primary bile acid CDCA, is identified in this study as a pathogenic factor contributing to intestinal barrier dysfunction in IBS-D. Moreover, LCA has been shown to induce apoptosis in the intestinal epithelium(*28*), which may further compromise barrier function in IBS-D. In contrast to the clinical findings in patients with IBS-D, LCA levels are significantly reduced in the serum and fecal samples of patients with IBD, according to clinical studies(*29, 30*). Given the reported anti-inflammatory effects of LCA in experimental colitis(*31*), the reduction of LCA in IBD may promote pro-inflammatory responses in IBD and subsequently impair barrier permeability. Therefore, manipulating bile acid homeostasis is crucial for managing intestinal barrier function in both IBS and IBD prior to the onset of symptoms.

In addition to the pathogenic role of LCA in IBS-D identified in this study, increased levels of LCA have been associated with various diseases. For example, excessive LCA is hepatotoxic and carcinogenic, damaging DNA and inducing colonic inflammation, which contributes to the development of colorectal cancer(*32*). Through the gut-brain axis, elevated LCA may also play a role in the pathogenesis of neurodegenerative diseases, such as Alzheimer’s and Parkinson’s disease, as well as psychological diseases like depression(*33–35*). Despite there are limited correlation and functional studies of LCA on brain function, excessive LCA in IBS may induce mental disorders by its biological roles in the enteric nervous system (ENS), inflammation and metabolism. However, correlation data of LCA with these diseases does not necessarily imply causation or demonstrate whether LCA contributes to the pathology of these diseases, so the exact role that LCA plays in these diseases remains to be investigated.

To date, there is no U.S. FDA-approved BA-targeted therapy for IBS and IBD, but bile acids sequestrants including cholestyramine, colestipol, and colesevelam are used to prevent bile acids diarrhea through binding bile acids in the gut(*36*). Our study highlights that BA-targeted therapy is promising for the drug development of both IBS and IBD to maintain barrier function and immune homeostasis. Activated by LCA, LPCAT1 is also a potential drug target that was implicated in the inflammatory processes of various GI diseases including colorectal cancer and IBD(*37*). Collectively, both LCA and LPCAT1-targeted therapeutics such as TSI-01 for the regulation of intestinal barrier function will be further studied. Our study also highlights the importance of lipid metabolism in the regulation of extracellular matrix dynamics and provides a potential therapeutic target LPCAT1 for diseases associated with MMP1 dysregulation.

A limitation of this study is that mice do not produce chenodeoxycholic acid, the precursor to LCA, which prevented us from investigating the effects of LCA-producing bacteria from human subjects in the recipient mice. This limitation hinders our ability to conclusively determine the causal relationship between LCA-producing bacteria and intestinal barrier dysfunction. Given this condition, future longitudinal studies and clinical trials targeting the LCA-LPCAT1 axis involving larger cohorts of patients with IBS-D can help elucidate the long-term relationship between intestinal barrier dysfunction and BA signaling. Further mechanistic studies are also warranted to refine our understanding of the molecular signaling pathways involving LPCAT1 and how they are regulated by other BAs.

In conclusion, this present study demonstrated that excessive LCA induced by gut dysbiosis in IBS-D contributed to intestinal barrier dysfunction through an LPCAT1-MMP1 axis, which led to the development of GI symptoms and colonic inflammation. Inhibition of LPCAT1 effectively mitigated LCA-induced intestinal barrier dysfunction, GI symptoms and colonic inflammation, offering a potential treatment strategy for IBS-D and IBD. These findings underscore the importance of host-microbiota interactions in maintaining GI health, providing novel therapeutic approaches targeting intestinal barrier dysfunction in GI diseases.

## Methods

### Reagents

Reagents and resource details are provided in Table.S1.

### Human study

#### Participants and recruitment

This study was approved by the Ethics Committee on the Use of Human & Animal Subjects in Teaching & Research of HKBU (Approval no. HASC/15-16/0300 and HASC/16-17/0027) and registered at clinicaltrials.gov (NCT02822677). The recruitment of participants and the collection of biological samples were conducted as described in a previous study(*6*). Specifically, adults aged 18-65 who met the Rome IV criteria for IBS-D were recruited from Chinese medicine clinics affiliated with the School of Chinese Medicine, Hong Kong Baptist University. Additionally, healthy volunteers, matched for age and sex, without gastrointestinal diseases, biliary-related diseases, or surgical histories involving gallbladder removal, GI tract, or cranial surgeries, were also included. All participants provided written informed consent for the use of their biological samples for scientific research.

#### Sample size

The present human study comprised a total sample size of 325 participants, consisting of 79 healthy volunteers and 246 patients diagnosed with IBS-D. The sample size calculation was derived from an observational cohort containing 30 participants, with a power of 80% and a two-tailed significance level of 5%.

#### Metabolomics analysis

For the serum samples, 100 μL of each serum sample was combined with 400 μL of the internal standard solution in methanol (50 nM CA-d4 and 50 nM dCA-d4) and mixed vigorously using a vortex mixer. For the tissue samples, about 100 mg of each tissue sample was extracted with 10x volume of 70% MeOH (m/v) combined with internal standard (50 nM CA-d4 and 50 nM dCA-d4) and then homogenized with steel beads. The samples were then centrifuged at 13,500 rpm for 15 minutes at 4°C. Subsequently, 200 μL of the supernatants were transferred into a liquid vial for LC-MS/MS analysis of bile acids profiles following the protocol established in our previous study(*6*).

#### Measurement of serum LBP and I-FABP

The serum levels of lipopolysaccharide-binding protein (LBP) and intestinal fatty acid-binding protein (I-FABP) were evaluated in 25 healthy controls and 83 patients with IBS-D. The serum levels of LBP and I-FABP were determined using commercial enzyme-linked immunosorbent assay (ELISA) kits. The serum concentrations of LBP and I-FABP were calculated by interpolating from their respective standard curves.

### Mouse study

Mouse studies were approved by the Hong Kong Baptist University Research Ethics Committee (No. REC/23-24/0457) and conducted following the regulations of the Animals (Control of Experiments) Ordinance of the Department of Health, Hong Kong SAR, China. Male C57BL/6J mice, aged 6-8 weeks and weighing 20-25 g, were purchased from the Laboratory Animal Services Centre, The Chinese University of Hong Kong (Hong Kong SAR, China). The mice were accommodated under a 12 h light/dark cycle at a regulated temperature of approximately 25°C and 60% humidity, with unrestricted access to food and water. The animal studies were conducted and reported following the ARRIVE guidelines(*38*).

#### Experiment 1: Treatment of LCA in normal and DSS-treated mice

The mouse model of experimental colitis was established by administering 2% dextran sulfate sodium in drinking water from Day 1 to Day 5. To investigate the action of LCA on intestinal barrier function and colonic inflammation in this model, mice were randomly assigned into 4 groups: Control group (n=6), which received a vehicle treatment from Day 1 to Day 12; LCA group (n=6), which received LCA (25 mg/kg) from Day 1 to Day 12; DSS group (n=6), which received 2% DSS in their drinking water from Day 1 to Day 5, followed by a vehicle treatment from Day 6 to Day 12; and DSS + LCA group (n=6), which received 2% DSS in their drinking water from Day 1 to Day 5, followed by LCA (25 mg/kg) from Day 6 to Day 12. Defection frequency and fecal water content were measured to determine the effects of LCA on GI motility.

#### Experiment 2: Generation of conditional knock in LPCAT1 ^flox/-^ mice

Mice were generated by Cyagen Biosciences (www.cyagen.com) using CRISPR/Cas9. The pathogenic variant was created in exon 3 of Fam161a using the following guide RNAs (gRNA): gRNA1 (matches the forward strand): CCACCGCGTCTTCCCGAGGGCGG, and gRNA2 (matches the reverse strand): GATGGCTTGCTCCCGCCCTCGGG (available at https://www.ophthalmologyscience.org). Potential off-target sites were analyzed. Off-target analysis for gRNA1 revealed 63 sites, 21 of which are located in genes. The genes with the highest probability of being affected by CRISPR/Cas9 manipulation are Xrn2, Crlf1, Sptbn4, Lbx2, Sec31b, Ogfod3, Gimap8, Scpep1os, and Nle1. Off-target analysis for gRNA2 revealed 116 sites, with 21 in genes. The genes with the highest probability of being affected by CRISPR/Cas9 manipulation are Thbd, Lrrc52, Ptprs, Syt16, Mroh8, Dot1l, Ncoa6, and Ptk2. None of those genes are expected to cause inherited retinal dystrophy (IRD). The guide RNA targeting vector and donor oligo (containing the targeting sequence flanked by 130 bp homologous sequences on both sides) were designed. The p.Arg512∗ (CGG to TGA) mutation sites in donor oligonucleotide were introduced into exon 3 by homology-directed repair. Cas9 messenger RNA (mRNA), *in vitro* transcribed gRNA, and donor oligonucleotide were co-injected into fertilized eggs of C57BL/6J mice to produce the knock-in mice.

8-10 week-old Rosa26-LSL-LPCAT1^flox/-^ mice (CKI mice) and villin-Cre LSL-LPCAT1^flox/flox^ mice (CRE mice) were administered a 2.0% (w/v) DSS solution via drinking water for 7 days to induce acute colitis as previously described(*39*). Both the DAI and histological scores were evaluated as described in our previous study(*40*) to determine the regulatory role of LPCAT1 on intestinal barrier function and inflammation in DSS-induced colitis mice.

#### Experiment 3: Inhibition of LPCAT1 in LCA-treated mice and DSS-induced colitis mice

A LPCAT1 inhibitor TSI-01 (10 mg/kg, i.p.) was administered concurrently with the DSS for a total duration of 7 days. Both the DAI and the histological scores were evaluated to determine the regulatory role of LPCAT1 on intestinal barrier function and inflammation in DSS-induced colitis mice. Subsequently, TSI-01 (10 mg/kg, i.p.) was administered concurrently with the LCA for a total duration of 7 days. Defection frequency and fecal water content were measured to determine the effects of LPCAT1 inhibitor on GI motility and intestinal barrier function in LCA-treated mice.

#### Evaluation of disease activity index (DAI)

For animals treated with DSS, colitis severity was assessed on the designated day by evaluating body weight, stool consistency, and fecal occult blood. The DAI scores were calculated based on weight loss, stool consistency, and bloody stool according to a standard protocol in our previous study(*40*). Fecal occult blood was determined using a Luminol reaction experiment kit. In a 96-well plate, 5 μL of feces supernatant was mixed with 5 μL of the 2-fold luminal reaction solution, and fluorescence was measured at 425 nm using a Spectra Max iD5(*41*).

#### Evaluation of intestinal barrier function

Mice were fasted for 3 h and orally administered a 750 mg/kg FITC-Dextran 4 kDa solution (750 mg in 10 mL of distilled water, 200 μL per mouse) to measure intestinal barrier function. After 4 hours, about 100 µL of blood was collected and allowed to stand for 30 min at room temperature, followed by centrifugation at 3000 rpm for 10 min. The supernatant was diluted with distilled water in a 1:4 ratio. All serum and standard curve samples were added to a 96-well plate for measurement. The fluorescence was measured at 485/525 nm.

#### Evaluation of transepithelial electrical resistance (TEER)

Mice were euthanized and sacrificed 12 days after treatment. Colon tissues were immediately collected, washed with Krebs-Ringer (KR) buffer and dissected longitudinally into small 3 ×1 cm²pieces. The colonic samples were incubated in Krebs-Ringer buffer for 10 min before being mounted between the two halves of the Ussing chamber. The Ussing chamber system was maintained at 37°C and provided with a carbogen (95% O₂, 5% CO₂) gas flow. After a 30-minute equilibration period, the solutions were replaced with fresh Krebs-Ringer buffer, and the experiments were initiated to collect TEER data within 30 min(*42*).

#### Histopathological analysis

Both hematoxylin and eosin (H&E) and Alcian blue staining were used to analyze the histopathological changes in colon tissues. Colon tissue samples were fixed in a 4% paraformaldehyde solution for 24 hours and subsequently dehydrated in sucrose at room temperature. After dehydration, the tissues were embedded in the O.C.T. compound upon collection. Frozen sections, with a thickness of 8 μm, underwent a 10-minute fixation in methanol. The resulting slices were then stained with Hematoxylin and Eosin (H&E) and Alcian Blue. Morphological evaluation and scoring of the tissues were conducted using an optical microscope, in accordance with the protocols established in our studies(*43, 44*).

### *In vitro* studies

#### Caco-2 cell culture

Caco-2 cells were cultured in 96-well plates at a density of 3 x 10³cells per well for 24 h. After treatment with LCA at different concentrations (0, 1, 10, 25, 50, 100, 250 μM) for 24 h, the cells were incubated with CCK8 for 3 h to measure cell viability via fluorescence at 450 nm.

#### Cell viability

Caco-2 cells were cultured in 96-well plates at a density of 3 x 10³cells/well for 24 h. After treatment of LCA at different concentrations (0, 1, 10, 25, 50, 100, 250 μM) for 24 h, the cells were incubated with CCK8 for 3 h for the measurement of cell viability through fluorescence at 450 nm.

#### Wound healing assay

Both LPCAT1-overexpressing Caco-2 cells and wildtype Caco-2 cells were seeded into 12-well plates and allowed to form a confluent monolayer for scratch assays. Caco-2 cells were scratched and rinsed with PBS, and the medium was replaced with 1% FBS medium. Images were captured using a microscope. Following imaging, treatment of TNF-α (100 ng/mL) or control was added for 24 hours and images were taken again to determine the wound healing ability. The scratch areas at 0 hours and 24 hours were measured, and the wound closure rate was calculated using the formula: (scratch area at 0 hours - scratch area at 24 hours) / scratch area at 0 hours ×100%.

#### Monolayer permeability assay

LPCAT1-overexpressing Caco-2 cells and wildtype Caco-2 cells were seeded into Transwell chambers for 28 days to evaluate cell permeability using the FITC-dextran, with fluorescence measured at 485/525 nm(*45*).

#### Evaluation of transepithelial electrical resistance (TEER)

Caco-2 cells were cultured in 12-well cell culture plates at a density of 6 x 10⁴ cells per well. The medium remained unchanged for the first three days and was then replaced daily until a single-cell layer barrier was formed (approximately 21 days). On day 21, LCA in different concentrations (0, 1, 10, 100 μM) were added. The TEER data of each group was measured 24 hours later using an electrical resistance system(*46*).

#### Lipidomics analysis

Caco-2 cells were cultured in 15cm dishes at a density of 3 x 10^7^ cells per dish for lipidomics analysis. During harvest, Caco-2 cells were prepared at a concentration of 6 x 10^6^ cells/mL, and washed twice with 5 mL cold 1x PBS, and then scraped and transferred to 15 mL tubes. Caco-2 cells were centrifuged at 3500 rpm for 10 min at 4°C, and the resulting pellet was resuspended in a mixture of pre-cooled Bligh and Dyer (BD) extraction system consisting of 700 μL MeOH, 280 μL H_2_O, and 350 μL CHCl_3_. The mixture was vortexed thoroughly and then centrifuged at 4000 rpm for 30 min at 4°C. The lower layer was transferred to a new 1.5 mL tube and dried under vacuum for 30 min at room temperature.

For the lipidomics analysis of colon tissue, approximately 50 mg of colon tissue was weighed and homogenized in PBS at a 1:10 (m/v) ratio. A 60 μL aliquot of the homogenate was mixed with 240 μL of Folch solvent (containing an internal standard) for lipid extraction. The mixture was vortexed and centrifuged at 8,000 rpm for 15 minutes at 4°C to achieve phase separation. Approximately 150 μL of the lower phase was collected and dried using a vacuum concentrator at room temperature within 30 minutes.

The resulting residue was dissolved in 200 μL of ACN: IPA: H2O (65:30:5, v/v/v) and subjected to analysis by LC-QTOF-MS.

#### Real-time quantitative PCR

Total RNA was isolated from either cells or frozen colonic tissues using RNAisoPlus. The RNA samples were then reverse-transcribed into cDNA using the PrimeScript RT Master Mix. Quantitative real-time PCR was performed using PowerTrack SYBR Green Master Mix. Primer lists of qPCR were provided in Table.S1.

#### Western blot

Proteins were extracted from either cells or frozen colonic tissues using RIPA lysis buffer. The extracted proteins were quantified using BCA protein assay kits. Equal amounts of protein (10 μg) were separated by 10% SDS-PAGE and transferred onto PVDF membranes. The membranes were blocked with 5% (w/v) BSA solution for 2 hours at room temperature, followed by overnight incubation at 4°C with primary antibodies. The membranes were then incubated with corresponding secondary antibodies for 2 hours at room temperature. The blots were developed using an ECL substrate and visualized using a gel imager. Densitometric analysis was performed using ImageJ software(*47*).

#### Thermal stability test of LPCAT1

Caco-2 cells were cultured in 6-well plates at a density of 9 × 10⁴ cells per well for 24 hours. For the thermal stability test, proteins from Caco-2 cells were extracted and divided into control and treatment groups as indicated. Proteins from all groups were incubated at room temperature for 1 hour. The extracts were then divided into six aliquots and heated at 40°C, 42°C, 44°C, 46°C, 50°C, and 58°C for 5 minutes each. Western blot analysis was performed to determine the protein expression levels of LPCAT1 under different temperature conditions.

#### Cycloheximide (CHX) chase assay

Caco-2 cells were cultured in 6-well plates at a density of 9 × 10⁴ cells per well for 24 hours. The medium was then replaced with a fresh complete medium containing 50 μg/mL CHX(*48*), followed by treatment with either 50 μM LCA or an equal volume of control medium. Proteins from Caco-2 cells were collected and extracted at 0, 2, 4, 8, 12, and 24 hours, respectively. Western blots were used to determine the protein expression levels of LPCAT1 at different time points.

#### Microscale thermophoresis (MST) analysis

Binding affinity measurements of purified LPCAT1 protein with various ligands were conducted using a Monolith NT.115 instrument(*49*). LPCAT1 was fluorescently labeled using the Monolith™ Series Protein Labeling Kit RED-NHS 2nd Generation. Ligands including LCA, deoxycholic acid (DCA) and chenodeoxycholic acid (CDCA) were prepared in a 16-step two-fold dilution series in PBST buffer. The diluted ligands were incubated with 2.5 μM labeled LPCAT1 at room temperature for 30 minutes, followed by centrifugation at 15,000 g for 5 minutes at 4°C to remove unbound ligands. The MST assay was performed using standard-treated capillaries, and data analysis was conducted with NanoTemper software, enabling precise quantification of binding affinities and insights into ligand-receptor interactions.

#### Ligand docking analysis

Molecular docking was conducted to analyze the binding affinity and interaction sites between LCA and LPCAT1. The structure of LCA was obtained from the PubChem database (CID 9903), while the LPCAT1 structure was obtained from the UniProt (Q8NF37). The results of the docking analysis were visualized using PyMOL (v2.2.0)(*50*).

#### Statistical methods

All data are presented as the mean ±standard deviation (SD) or standard error of mean (SEM) based on a minimum of three independent experiments. Statistical analyses were conducted using GraphPad Prism software 10. One-way or two-way analysis of variance (ANOVA) was used to assess statistical differences among three or more groups. A *p*-value of less than 0.05 was considered statistically significant. The Kruskal-Wallis test was used to compare multiple groups for variations in clinical characteristics, bowel symptom severity scores, and BA-related indices. The Mann-Whitney test was employed for comparisons between the two groups. Additionally, the Benjamin-Hochberg method was used to identify differential taxa and BA-transforming genomes, with a significance threshold of p < 0.1. Basic research studies did not apply statistical methods to establish sample sizes beforehand, and all data collected were included in the analysis. The study design did not involve randomization of experiments, and the investigators were aware of the allocation during the experiments and when assessing outcomes.

## Data Availability

All data produced in the present study are available upon reasonable request to the authors

## Authorship Contributions

L.Z and Z.B are the principal investigators and design this study. L.Z. and H.X. drafted the manuscript. H.X., B.L., J.P., Y.Z., J.H. and H.D. conduct the *in vivo* and *in vitro* studies including animal experiments and cell experiments. S.X. and G.B. conducted the metabolomics analysis and lipidomics analysis. G.B. and Y.L. manage the clinical specimens collected from all participants. X.HL.W, L.Z and C.L. provided important suggestions towards the study design and helped revise the manuscript. All authors have read and approved the manuscript.

## Acknowledgments

The authors acknowledge all the participants in this study. This study is supported by the General Research Fund (12100323 to Zhaoxiang Bian) and Health@InnoHK Initiative Fund from the Hong Kong SAR Government (ITC RC/IHK/4/7 to Zhaoxiang Bian), the Guangdong Basic and Applied Basic Research Foundation (2414050003325 to Lixiang Zhai), Guangdong Natural Science Foundation (2023A1515012475 to Haitao Xiao), Foundation of Shenzhen Science and Technology Innovation Committee (JCYJ20230807095119038 to Jiao Peng) and the National Natural Science Foundation of China (82370553 to Ling Zhao). The authors also thank the Vincent and Lily Woo Foundation for their support of this work. The authors declare that they have no competing interests. All authors read and approved the manuscript.

## Abbreviations

BAs: Bile acids
DSS: Dextran sulfate sodium
IBS-D: Diarrhea-predominant Irritable bowel syndrome
DAI: Disease activity index
ENS: Enteric nervous system
FD4: Fluorescein isothiocyanate-dextran
GI: Gastrointestinal
GEO: Gene Expression Omnibus
GM: Germ-free
HC: Healthy controls
H&E: Hematoxylin and eosin
IBD: Inflammatory bowel disease
I-FABP: Intestinal fatty acid binding protein
IBS: Irritable bowel syndrome
LBP: Lipopolysaccharide binding protein
LCA: Lithocholic acid
LysoPC: Lysophosphatidylcholine
LPCAT1: Lysophosphatidylcholine acyltransferase 1
MMP1: Matrix metallopeptidase 1
MMP1: Matrix metalloproteinase 1
PC: Phosphatidylcholine
SBA: Secondary bile acids
SPF: Specific pathogen-free
TEER: Transepithelial electrical resistance
UC: Ulcerative colitis
WT: Wide type

## References

1. R. Spiller, G. Major, IBS and IBD - separate entities or on a spectrum? Nat Rev Gastroenterol Hepatol 13, 613–621 (2016).

2. C. K. Porter, B. D. Cash, M. Pimentel, A. Akinseye, M. S. Riddle, Risk of inflammatory bowel disease following a diagnosis of irritable bowel syndrome. BMC Gastroenterology 12, 55 (2012).

3. M. Tena-Garitaonaindia, M. Arredondo-Amador, C. Mascaraque, M. Asensio, J. J. G. Marin, O. Martínez-Augustin, F. Sánchez de Medina, Modulation of intestinal barrier function by glucocorticoids: Lessons from preclinical models. Pharmacol Res 177, 106056 (2022).

4. J. König, J. Wells, P. D. Cani, C. L. García-Ródenas, T. MacDonald, A. Mercenier, J. Whyte, F. Troost, R. J. Brummer, Human Intestinal Barrier Function in Health and Disease. Clin Transl Gastroenterol 7, e196 (2016).

5. C. Casén, H. C. Vebø, M. Sekelja, F. T. Hegge, M. K. Karlsson, E. Ciemniejewska, S. Dzankovic, C. Frøyland, R. Nestestog, L. Engstrand, P. Munkholm, O. H. Nielsen, G. Rogler, M. Simrén, L. Öhman, M. H. Vatn, K. Rudi, Deviations in human gut microbiota: a novel diagnostic test for determining dysbiosis in patients with IBS or IBD. Aliment Pharmacol Ther 42, 71–83 (2015).

6. L. Zhao, W. Yang, Y. Chen, F. Huang, L. Lu, C. Lin, T. Huang, Z. Ning, L. Zhai, L. L. Zhong, W. Lam, Z. Yang, X. Zhang, C. Cheng, L. Han, Q. Qiu, X. Shang, R. Huang, H. Xiao, Z. Ren, D. Chen, S. Sun, H. El-Nezami, Z. Cai, A. Lu, X. Fang, W. Jia, Z. Bian, A Clostridia-rich microbiota enhances bile acid excretion in diarrhea-predominant irritable bowel syndrome. J Clin Invest 130, 438–450 (2020).

7. O. Chávez-Talavera, A. Tailleux, P. Lefebvre, B. Staels, Bile Acid Control of Metabolism and Inflammation in Obesity, Type 2 Diabetes, Dyslipidemia, and Nonalcoholic Fatty Liver Disease. Gastroenterology 152, 1679–1694.e1673 (2017).

8. B. van der Lugt, M. C. P. Vos, M. Grootte Bromhaar, N. Ijssennagger, F. Vrieling, J. Meijerink, W. T. Steegenga, The effects of sulfated secondary bile acids on intestinal barrier function and immune response in an inflammatory in vitro human intestinal model. Heliyon 8, e08883 (2022).

9. N. L.-M. Ginley, J. Smyth, C. Fallon, C. Clerkin, S. J. Keely, Lithocholic acid regulates colonic epithelial barrier function and apoptosis: Implications for inflammatory bowel disease. The FASEB Journal 33, 869.817–869.817 (2019).

10. M. Linsalata, G. Riezzo, C. Clemente, B. D’Attoma, F. Russo, Noninvasive Biomarkers of Gut Barrier Function in Patients Suffering from Diarrhea Predominant-IBS: An Update. Dis Markers 2020, 2886268 (2020).

11. L. Han, L. Zhao, Y. Zhou, C. Yang, T. Xiong, L. Lu, Y. Deng, W. Luo, Y. Chen, Q. Qiu, X. Shang, L. Huang, Z. Mo, S. Huang, S. Huang, Z. Liu, W. Yang, L. Zhai, Z. Ning, C. Lin, T. Huang, C. Cheng, L. L. D. Zhong, S. Li, Z. Bian, X. Fang, Altered metabolome and microbiome features provide clues in understanding irritable bowel syndrome and depression comorbidity. Isme j 16, 983–996 (2022).

12. S. Carlin, J. P. Kennelly, H. Fedoruk, A. Quiroga, K. A. Leonard, R. Nelson, A. Thiesen, J. Buteau, R. Lehner, R. Jacobs, De novo phosphatidylcholine synthesis in the small intestinal epithelium is required for normal dietary lipid handling and maintenance of the mucosal barrier. Biochim Biophys Acta Mol Cell Biol Lipids 1867, 159109 (2022).

13. R. Ehehalt, A. Braun, M. Karner, J. Füllekrug, W. Stremmel, Phosphatidylcholine as a constituent in the colonic mucosal barrier--physiological and clinical relevance. Biochim Biophys Acta 1801, 983–993 (2010).

14. Z. Li, Y. Hu, H. Zheng, M. Li, Y. Liu, R. Feng, X. Li, S. Zhang, M. Tang, M. Yang, R. Yu, Y. Xu, X. Liao, S. Chen, W. Qian, Q. Zhang, D. Tang, B. Li, L. Song, J. Li, LPCAT1-mediated membrane phospholipid remodelling promotes ferroptosis evasion and tumour growth. Nat Cell Biol 26, 811–824 (2024).

15. V. Mariaule, A. Kriaa, S. Soussou, S. Rhimi, H. Boudaya, J. Hernandez, E. Maguin, A. Lesner, M. Rhimi, Digestive Inflammation: Role of Proteolytic Dysregulation. Int J Mol Sci 22, (2021).

16. R. E. Vandenbroucke, C. Libert, Is there new hope for therapeutic matrix metalloproteinase inhibition? Nat Rev Drug Discov 13, 904–927 (2014).

17. Y. D. Wang, P. Y. Yan, Expression of matrix metalloproteinase-1 and tissue inhibitor of metalloproteinase-1 in ulcerative colitis. World J Gastroenterol 12, 6050–6053 (2006).

18. K. Kobayashi, Y. Arimura, A. Goto, S. Okahara, T. Endo, Y. Shinomura, K. Imai, Therapeutic implications of the specific inhibition of causative matrix metalloproteinases in experimental colitis induced by dextran sulphate sodium. J Pathol 209, 376–383 (2006).

19. F. Wu, T. Dassopoulos, L. Cope, A. Maitra, S. R. Brant, M. L. Harris, T. M. Bayless, G. Parmigiani, S. Chakravarti, Genome-wide gene expression differences in Crohn’s disease and ulcerative colitis from endoscopic pinch biopsies: insights into distinctive pathogenesis. Inflamm Bowel Dis 13, 807–821 (2007).

20. J. Olsen, T. A. Gerds, J. B. Seidelin, C. Csillag, J. T. Bjerrum, J. T. Troelsen, O. H. Nielsen, Diagnosis of ulcerative colitis before onset of inflammation by multivariate modeling of genome-wide gene expression data. Inflamm Bowel Dis 15, 1032–1038 (2009).

21. R. Häsler, Z. Feng, L. Bäckdahl, M. E. Spehlmann, A. Franke, A. Teschendorff, V. K. Rakyan, T. A. Down, G. A. Wilson, A. Feber, S. Beck, S. Schreiber, P. Rosenstiel, A functional methylome map of ulcerative colitis. Genome Res 22, 2130–2137 (2012).

22. N. Planell, J. J. Lozano, R. Mora-Buch, M. C. Masamunt, M. Jimeno, I. Ordás, M. Esteller, E. Ricart, J. M. Piqué, J. Panés, A. Salas, Transcriptional analysis of the intestinal mucosa of patients with ulcerative colitis in remission reveals lasting epithelial cell alterations. Gut 62, 967–976 (2013).

23. A. Acciarino, S. Diwakarla, J. Handreck, C. Bergola, L. Sahakian, R. M. McQuade, The role of the gastrointestinal barrier in obesity-associated systemic inflammation. Obes Rev 25, e13673 (2024).

24. N. Hanning, A. L. Edwinson, H. Ceuleers, S. A. Peters, J. G. De Man, L. C. Hassett, B. Y. De Winter, M. Grover, Intestinal barrier dysfunction in irritable bowel syndrome: a systematic review. Therapeutic advances in gastroenterology 14, 1756284821993586 (2021).

25. L. Zhai, C. Huang, Z. Ning, Y. Zhang, M. Zhuang, W. Yang, X. Wang, J. Wang, L. Zhang, H. Xiao, L. Zhao, P. Asthana, Y. Y. Lam, C. F. W. Chow, J. D. Huang, S. Yuan, K. M. Chan, C. S. Yuan, J. Y. Lau, H. L. X. Wong, Z. X. Bian, Ruminococcus gnavus plays a pathogenic role in diarrhea-predominant irritable bowel syndrome by increasing serotonin biosynthesis. Cell Host Microbe 31, 33–44.e35 (2023).

26. L. Zhao, W. Yang, Y. Chen, F. Huang, L. Lu, C. Lin, T. Huang, Z. Ning, L. Zhai, L. L. D. Zhong, W. Lam, Z. Yang, X. Zhang, C. Cheng, L. Han, Q. Qiu, X. Shang, R. Huang, H. Xiao, Z. Ren, D. Chen, S. Sun, H. El-Nezami, Z. Cai, A. Lu, X. Fang, W. Jia, Z. Bian, A Clostridia-rich microbiota enhances bile acid excretion in diarrhea-predominant irritable bowel syndrome. The Journal of Clinical Investigation 130, 438–450 (2024).

27. Y. Kinashi, K. Hase, Partners in Leaky Gut Syndrome: Intestinal Dysbiosis and Autoimmunity. Front Immunol 12, 673708 (2021).

28. Z. Chen, Y. Liu, Research progress on relationship between bile acids and intestinal mucosal mechanical barrier function. Chinese Journal of Digestive Surgery 17, 967–970 (2018).

29. R.-H. Xu, J.-N. Shen, J.-B. Lu, Y.-J. Liu, Y. Song, Y. Cao, Z.-H. Wang, J. Zhang, Bile acid profiles and classification model accuracy for inflammatory bowel disease diagnosis. Medicine 103, e38457 (2024).

30. E. A. Franzosa, A. Sirota-Madi, J. Avila-Pacheco, N. Fornelos, H. J. Haiser, S. Reinker, T. Vatanen, A. B. Hall, H. Mallick, L. J. McIver, J. S. Sauk, R. G. Wilson, B. W. Stevens, J. M. Scott, K. Pierce, A. A. Deik, K. Bullock, F. Imhann, J. A. Porter, A. Zhernakova, J. Fu, R. K. Weersma, C. Wijmenga, C. B. Clish, H. Vlamakis, C. Huttenhower, R. J. Xavier, Author Correction: Gut microbiome structure and metabolic activity in inflammatory bowel disease. Nat Microbiol 4, 898 (2019).

31. S. He, J. Li, Z. Yao, Z. Gao, Y. Jiang, X. Chen, L. Peng, Insulin alleviates murine colitis through microbiome alterations and bile acid metabolism. Journal of Translational Medicine 21, 498 (2023).

32. Q. Yang, B. Wang, Q. Zheng, H. Li, X. Meng, F. Zhou, L. Zhang, A Review of Gut Microbiota-Derived Metabolites in Tumor Progression and Cancer Therapy. Advanced Science 10, 2207366 (2023).

33. Z. Ren, L. Zhao, M. Zhao, T. Bao, T. Chen, A. Zhao, X. Zheng, X. Gu, T. Sun, Y. Guo, Y. Tang, G. Xie, W. Jia, Increased intestinal bile acid absorption contributes to age-related cognitive impairment. Cell Reports Medicine 5, (2024).

34. S. MahmoudianDehkordi, M. Arnold, K. Nho, S. Ahmad, W. Jia, G. Xie, G. Louie, A. Kueider-Paisley, M. A. Moseley, J. W. Thompson, L. St John Williams, J. D. Tenenbaum, C. Blach, R. Baillie, X. Han, S. Bhattacharyya, J. B. Toledo, S. Schafferer, S. Klein, T. Koal, S. L. Risacher, M. Allan Kling, A. Motsinger-Reif, D. M. Rotroff, J. Jack, T. Hankemeier, D. A. Bennett, P. L. De Jager, J. Q. Trojanowski, L. M. Shaw, M. W. Weiner, P. M. Doraiswamy, C. M. van Duijn, A. J. Saykin, G. Kastenmüller, R. Kaddurah-Daouk, f. t. A. s. D. N. Initiative, t. A. D. M. Consortium, Altered bile acid profile associates with cognitive impairment in Alzheimer’s disease—An emerging role for gut microbiome. Alzheimer’s & Dementia 15, 76–92 (2019).

35. H. Weng, L. Deng, T. Wang, H. Xu, J. Wu, Q. Zhou, L. Yu, B. Chen, L. a. Huang, Y. Qu, L. Zhou, X. Chen, Humid heat environment causes anxiety-like disorder via impairing gut microbiota and bile acid metabolism in mice. Nature Communications 15, 5697 (2024).

36. M. Camilleri, P. Vijayvargiya, The Role of Bile Acids in Chronic Diarrhea. Official journal of the American College of Gastroenterology | ACG 115, 1596–1603 (2020).

37. B. Wang, P. Tontonoz, Phospholipid Remodeling in Physiology and Disease. Annual Review of Physiology 81, 165–188 (2019).

38. N. Percie du Sert, V. Hurst, A. Ahluwalia, S. Alam, M. T. Avey, M. Baker, W. J. Browne, A. Clark, I. C. Cuthill, U. Dirnagl, M. Emerson, P. Garner, S. T. Holgate, D. W. Howells, N. A. Karp, S. E. Lazic, K. Lidster, C. J. MacCallum, M. Macleod, E. J. Pearl, O. H. Petersen, F. Rawle, P. Reynolds, K. Rooney, E. S. Sena, S. D. Silberberg, T. Steckler, H. Würbel, The ARRIVE guidelines 2.0: Updated guidelines for reporting animal research. PLoS Biol 18, e3000410 (2020).

39. S. Wirtz, V. Popp, M. Kindermann, K. Gerlach, B. Weigmann, S. Fichtner-Feigl, M. F. Neurath, Chemically induced mouse models of acute and chronic intestinal inflammation. Nat Protoc 12, 1295–1309 (2017).

40. L. Zhai, J. Peng, M. Zhuang, Y. Y. Chang, K. W. Cheng, Z. W. Ning, T. Huang, C. Lin, H. L. X. Wong, Y. Y. Lam, H. Y. Tan, H. T. Xiao, Z. X. Bian, Therapeutic effects and mechanisms of Zhen-Wu-Bu-Qi Decoction on dextran sulfate sodium-induced chronic colitis in mice assessed by multi-omics approaches. Phytomedicine 99, 154001 (2022).

41. A.-M. Park, I. Tsunoda, Forensic Luminol Reaction for Detecting Fecal Occult Blood in Experimental Mice. BioTechniques 65, 227–230 (2018).

42. J.-y. Zhou, W.-w. Xie, T.-c. Hu, X.-f. Wang, H.-c. Yan, X.-q. Wang, Mulberry Leaf-Derived Morin Activates β-Catenin by Binding to Frizzled7 to Promote Intestinal Stem Cell Expansion upon Heat-Stable Enterotoxin b Injury. Journal of Agricultural and Food Chemistry 72, 10366–10375 (2024).

43. Z. Wu, S. Huang, T. Li, N. Li, D. Han, B. Zhang, Z. Z. Xu, S. Zhang, J. Pang, S. Wang, G. Zhang, J. Zhao, J. Wang, Gut microbiota from green tea polyphenol-dosed mice improves intestinal epithelial homeostasis and ameliorates experimental colitis. Microbiome 9, 184 (2021).

44. L. Huang, J. Zheng, G. Sun, H. Yang, X. Sun, X. Yao, A. Lin, H. Liu, 5-Aminosalicylic acid ameliorates dextran sulfate sodium-induced colitis in mice by modulating gut microbiota and bile acid metabolism. Cellular and Molecular Life Sciences 79, 460 (2022).

45. A. Lauffer, T. Vanuytsel, F. Fornari, R. Farré, Su1870 Differential Effects of Acute, Chronic and Combined Stress Paradigms on Small Intestinal and Colonic Permeability in Rats. Gastroenterology 146, (2014).

46. S. Lopez-Escalera, A. Wellejus, Evaluation of Caco-2 and human intestinal epithelial cells as in vitro models of colonic and small intestinal integrity. Biochemistry and Biophysics Reports 31, 101314 (2022).

47. C. Wei, X. Dong, H. Lu, F. Tong, L. Chen, R. Zhang, J. Dong, Y. Hu, G. Wu, X. Dong, LPCAT1 promotes brain metastasis of lung adenocarcinoma by up-regulating PI3K/AKT/MYC pathway. Journal of Experimental & Clinical Cancer Research 38, 95 (2019).

48. H. G. Zhang, B. Wang, Y. Yang, X. Liu, J. Wang, N. Xin, S. Li, Y. Miao, Q. Wu, T. Guo, Y. Yuan, Y. Zuo, X. Chen, T. Ren, C. Dong, J. Wang, H. Ruan, M. Sun, X. Xu, H. Zheng, Depression compromises antiviral innate immunity via the AVP-AHI1-Tyk2 axis. Cell Res 32, 897–913 (2022).

49. L. Nie, Y. Xiao, T. Zhou, H. Feng, M. He, Q. Liang, K. Mu, H. Nie, Q. Huang, W. Chen, Cyclic di-GMP inhibits nitrate assimilation by impairing the antitermination function of NasT in Pseudomonas putida. Nucleic Acids Research 52, 186–203 (2023).

50. Y. Bi, S. Liu, X. Qin, M. Abudureyimu, L. Wang, R. Zou, A. Ajoolabady, W. Zhang, H. Peng, J. Ren, Y. Zhang, FUNDC1 interacts with GPx4 to govern hepatic ferroptosis and fibrotic injury through a mitophagy-dependent manner. Journal of Advanced Research 55, 45–60 (2024).

