## Supplemental Figures for "Gut microbiota-derived lithocholic acid leads to intestinal barrier dysfunction via LPCAT1 in irritable bowel syndrome"

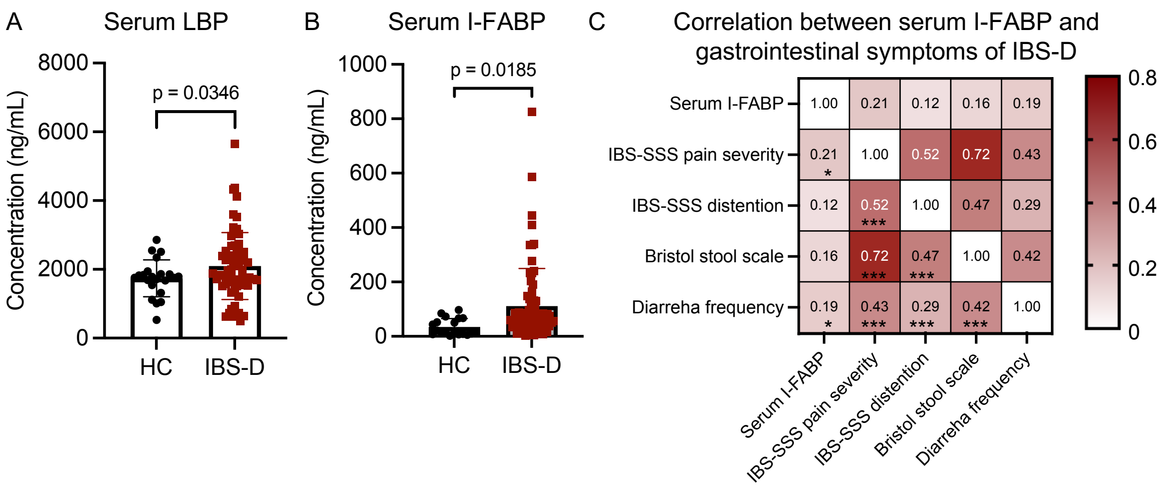


**Figure.S1 Intestinal barrier dysfunction is positively associated with GI symptoms in patients with IBS-D. (A-B)** Serum levels of LBP and I-FABP in patients with IBS-D (n=83) and healthy controls (HC, n=25). Data were analyzed using a two-tailed t-test. **(C)** Nonparametric spearman correlation analysis between serum I-FABP levels and the severity of GI symptoms in IBS-D patients (one-tailed).

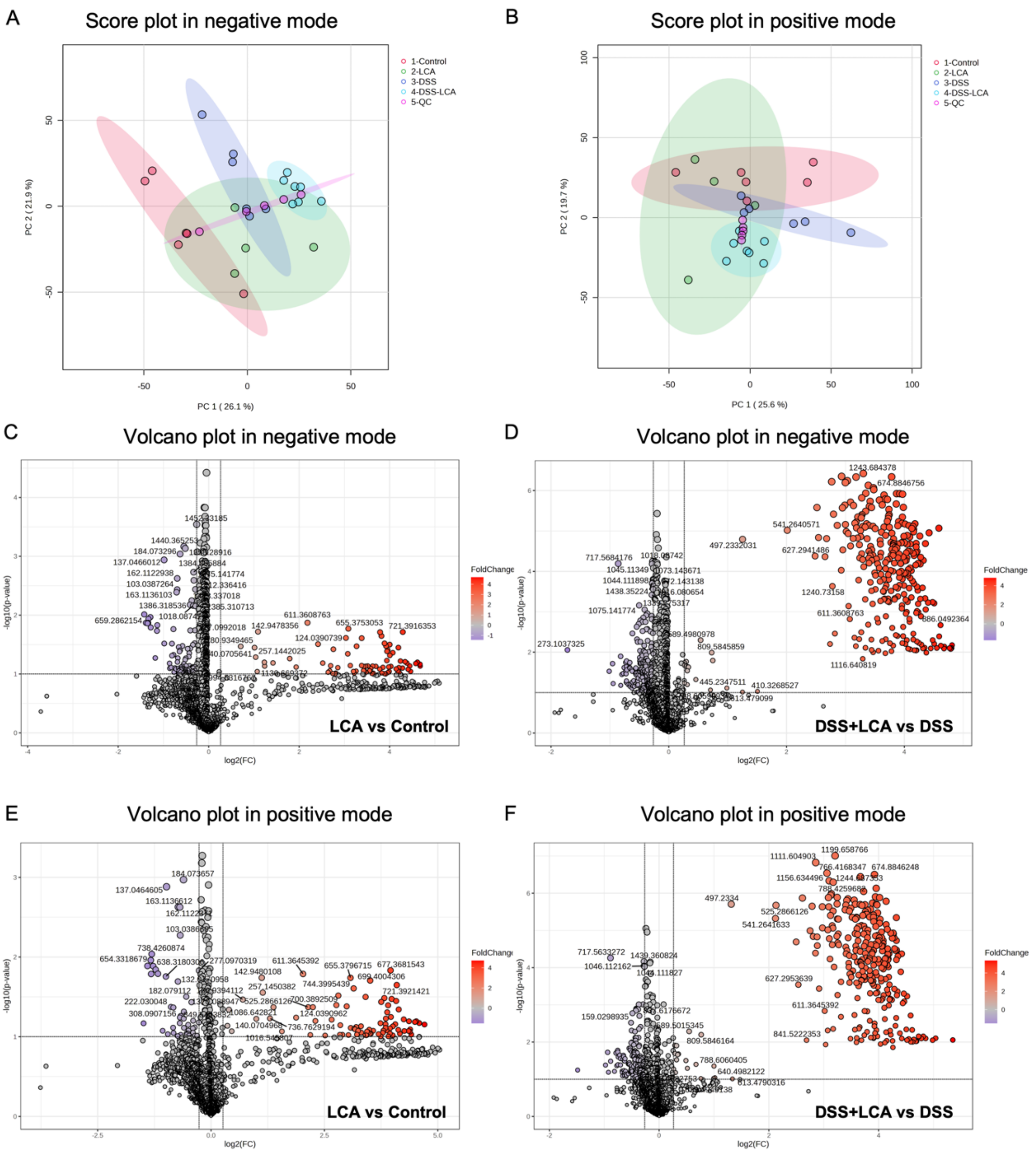


**Figure.S2 LCA alters phospholipid profiles in normal and DSS-treated mice.** (**A-B**) Score plot of principal component analysis on lipidome in colonic tissues from mice treated with vehicle control or LCA in both normal and inflammatory conditions in both negative and positive mode (n=6 per group). **(C-F)** Volcano plots of selected ions of lipidome with thresholds of fold change > 1.2 and p-value < 0.05 in colonic tissues from mice treated with vehicle control or LCA in both normal and inflammatory conditions in both negative and positive mode (n=6 per group).

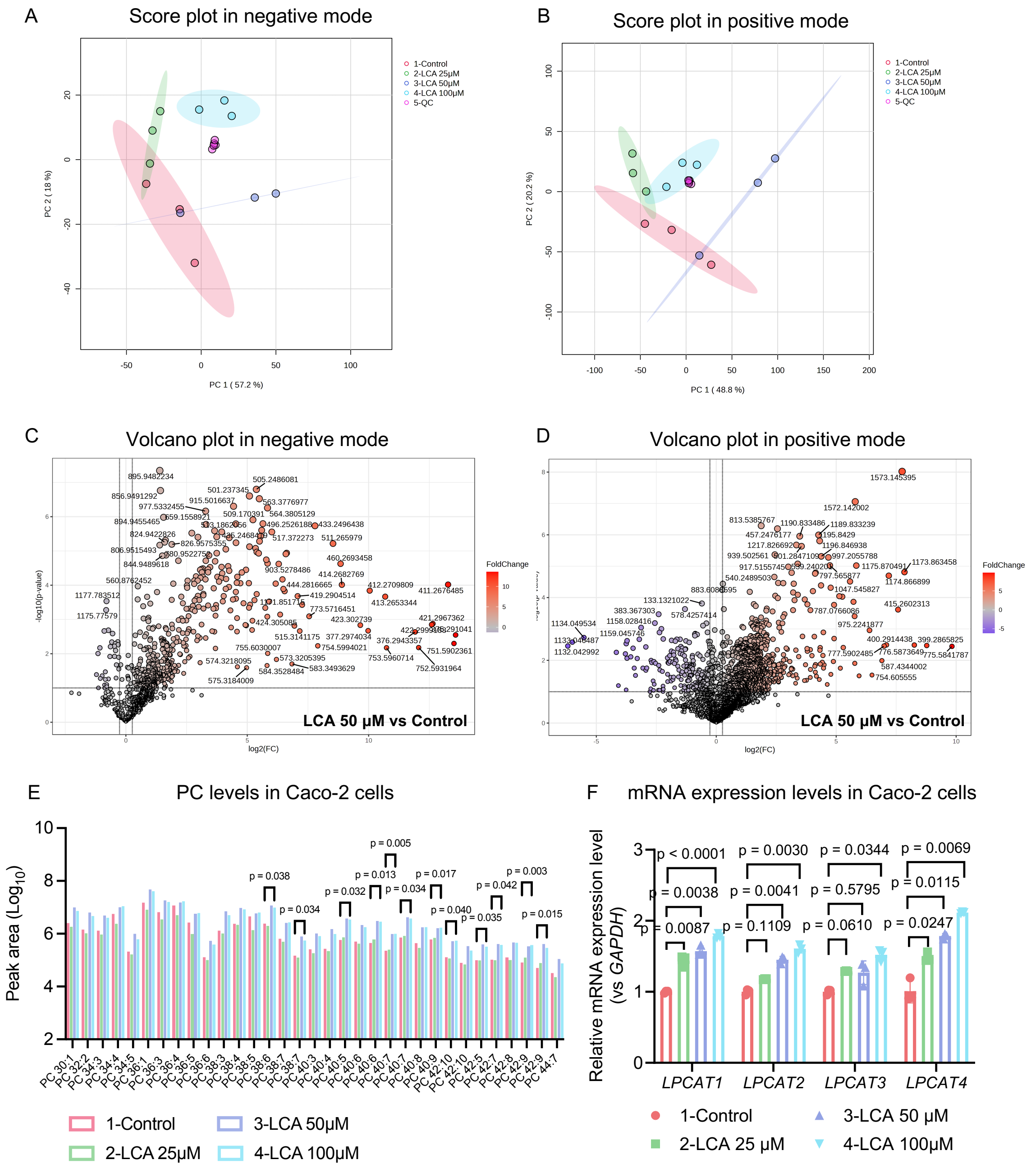


**Figure.S3 LCA upregulates PC levels and LPCAT1 expression in Caco-2 cells.** (**A-B**) Score plots of principal component analysis on lipidome in Caco-2 cells treated with vehicle control or LCA in both negative and positive mode (n=3 per group). **(C-D)** Volcano plots of selected lipidome with thresholds of fold change > 1.2 and p-value < 0.05 in Caco-2 cells treated with vehicle control or LCA in both negative and positive mode (n=3 per group). **(E)** PC levels in Caco-2 cells treated with vehicle control or LCA (n=3 per group). **(F)** mRNA expression of *LPCAT1, LPCAT2, LPCAT3* and *LPCAT4* levels in Caco-2 cells treated with vehicle control or LCA (n=3 per group). Data were analyzed using a two-tailed two-way ANOVA.

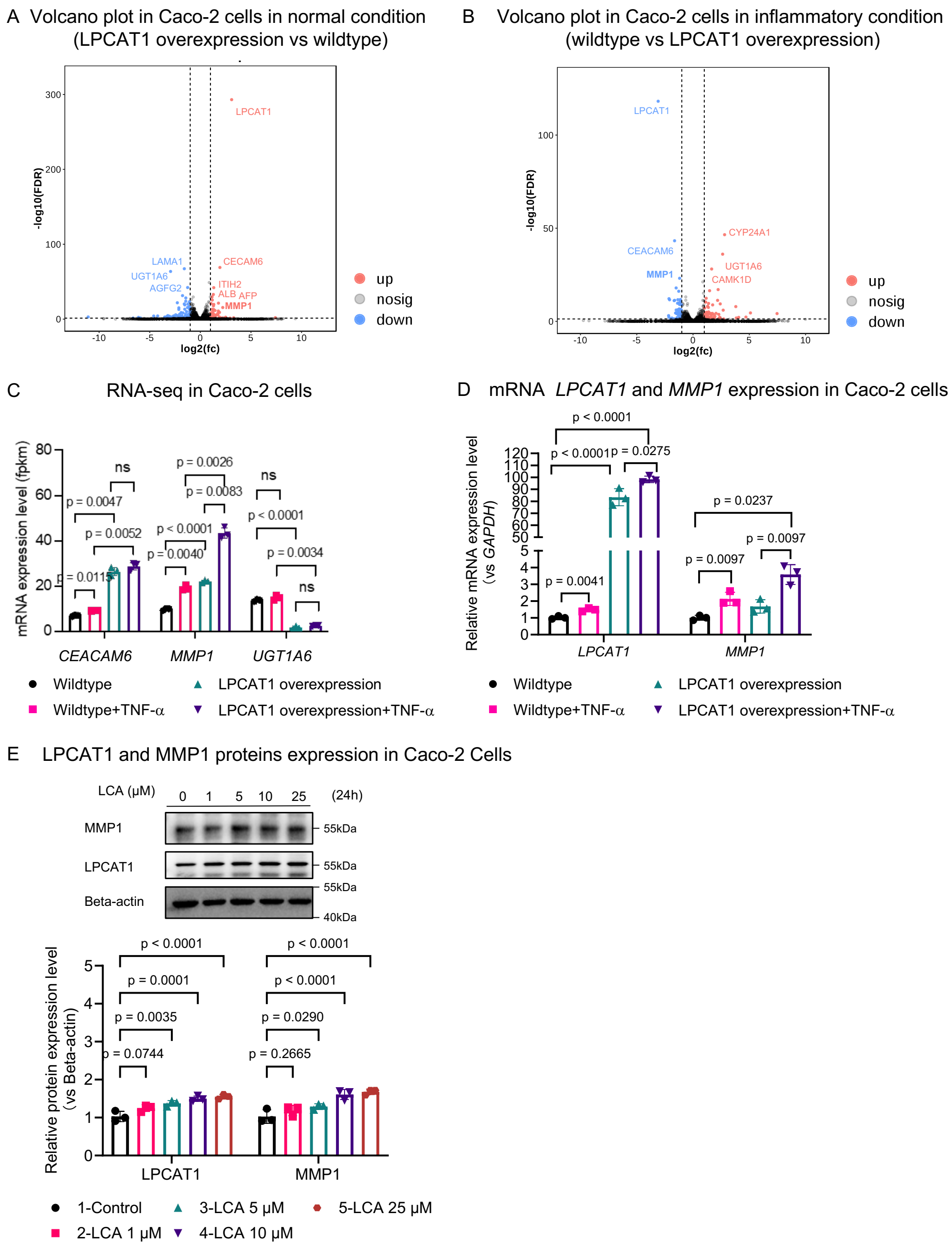


**Figure.S4 LPCAT1 upregulation by LCA induces pro-inflammatory protease MMP1 secretion in Caco-2 cells. (A-B)** Volcano plots in LPCAT1-overexpressed Caco-2 cells under normal and inflammatory conditions. **(C)** mRNA expression levels of *CEACAM6*, *MMP1* and *UGT1A6* in wildtype and LPCAT1-overexpressed Caco-2 cells in normal and inflammatory conditions measured by RNA-seq analysis (n=3 per group). **(D)** mRNA expression levels of *LPCAT1* and *MMP1* in wildtype and LPCAT1-overexpressed Caco-2 cells in normal and inflammatory conditions measured by qPCR analysis (n=3 per group). **(E)** Western blot analysis and semi-quantification of LPCAT1 and MMP1 in Caco-2 cells treated with vehicle control or LCA at indicated concentrations (n=3 per group). Data were analyzed using a two-tailed two-way ANOVA.

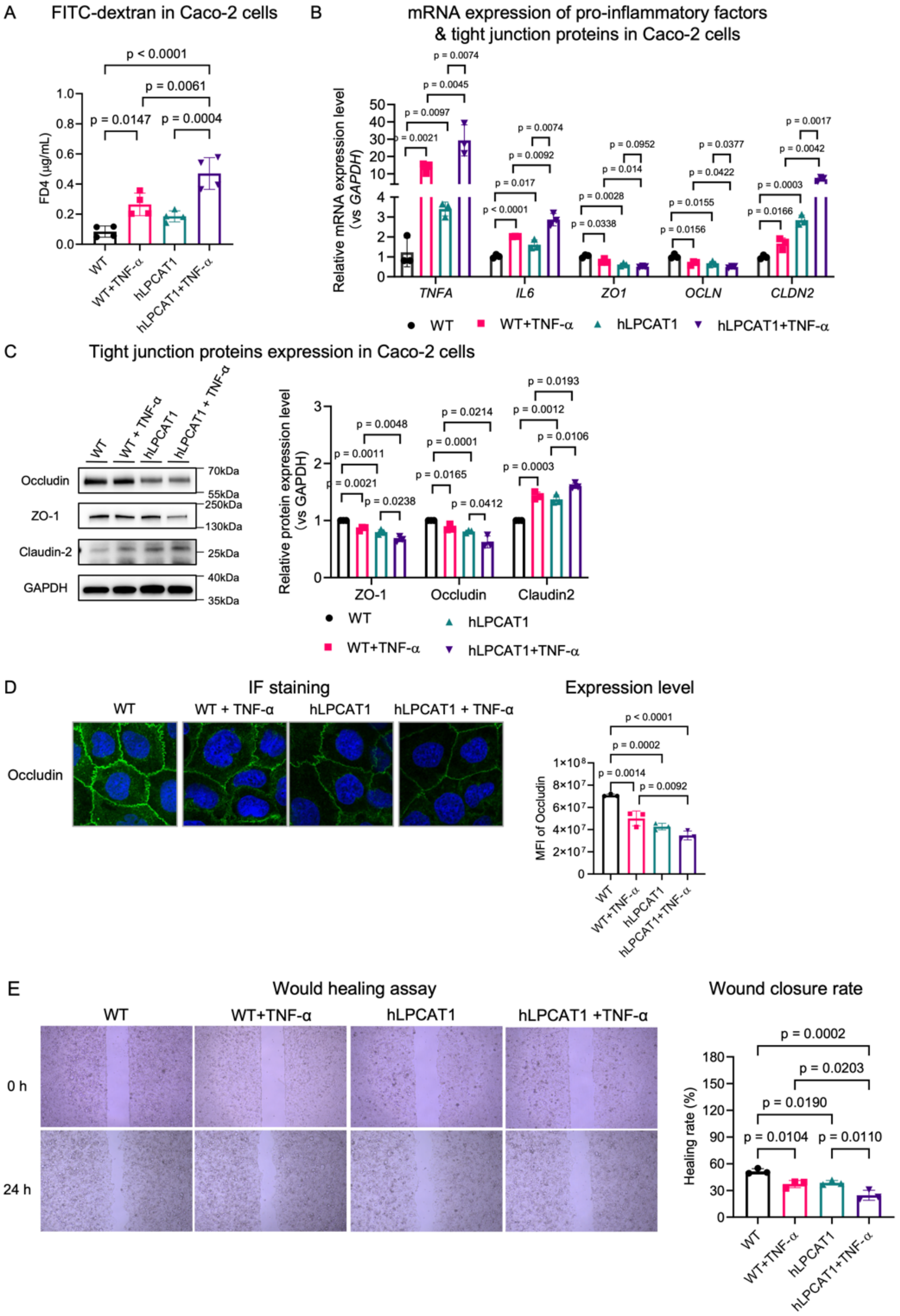


**Figure.S5 LPCAT1 overexpression impairs intestinal barrier integrity and induces pro-inflammatory responses in Caco-2 cells. (A)** FITC-Dextran in WT and LPCAT1 over-expressed Caco-2 cells treated with and without TNF-α (n=3 per group). **(B)** mRNA expression levels of pro-inflammatory factors and tight junction proteins in WT and LPCAT1 over-expressed Caco-2 cells treated with and without TNF-α (n=3 per group). **(C)** Western blot analysis and semi-quantification of ZO-1, Occludin and Claudin 2 in WT and LPCAT1 over-expressed Caco-2 cells treated with and without TNF-α (n=3 per group). **(D)** IF staining of Occludin in WT and LPCAT1 over-expressed Caco-2 cells treated with and without TNF-α (n=3 per group). **(E)** Wound healing closure rate in WT and LPCAT1 over-expressed Caco-2 cells treated with and without TNF-α (n=3 per group). Data were analyzed using a two-tailed one-way ANOVA.

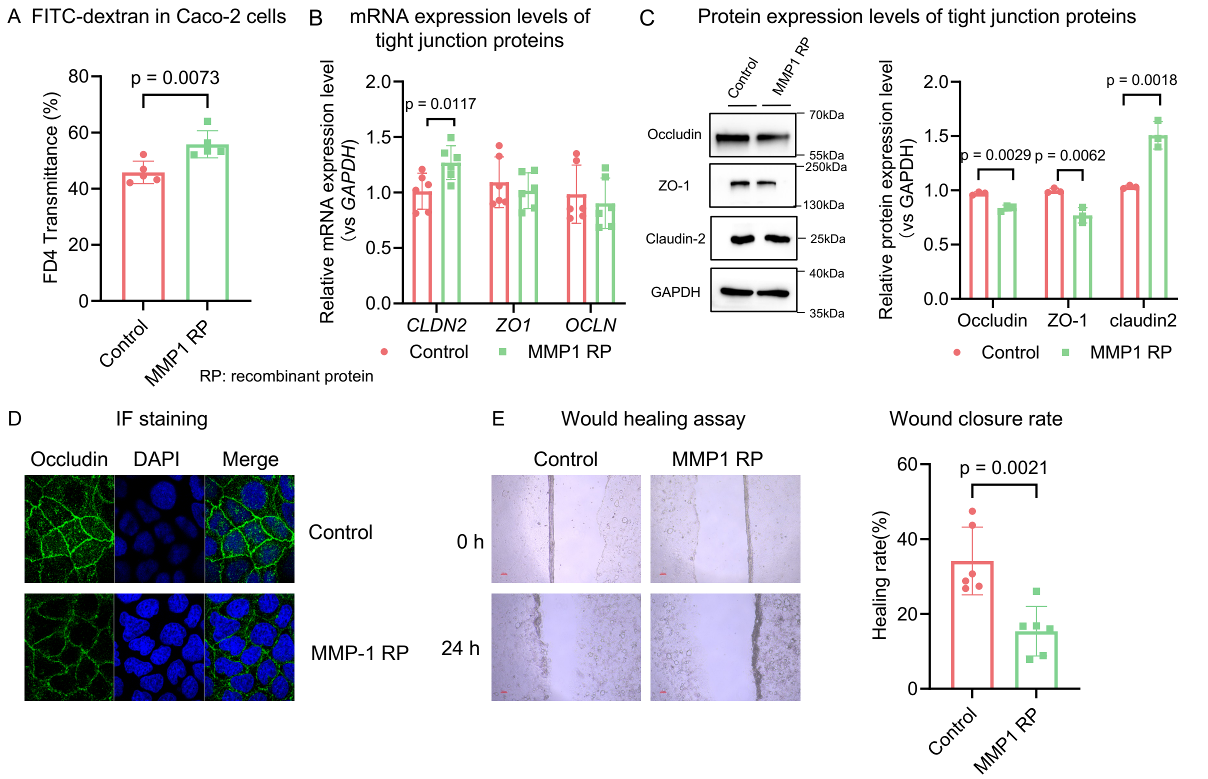


**Figure.S6 MMP1 recombinant protein disrupts intestinal barrier integrity in Caco-2 cells.** **(A-B)** FITC-Dextran and mRNA expression levels of tight junction proteins in Caco-2 cells treated with and without MMP1 recombinant protein (1 µM) (n=3 per group). **(C)** Western blot analysis and semi-quantification of ZO-1, Occludin and Claudin 2 in Caco-2 cells treated with and without MMP1 recombinant protein (1 µM) (n=3 per group). **(D-E)** IF staining of Occludin and wound healing closure rate in Caco-2 cells treated with and without MMP1 recombinant protein (1 µM) (n=3 per group). Data were analyzed using a two-tailed one-way ANOVA.

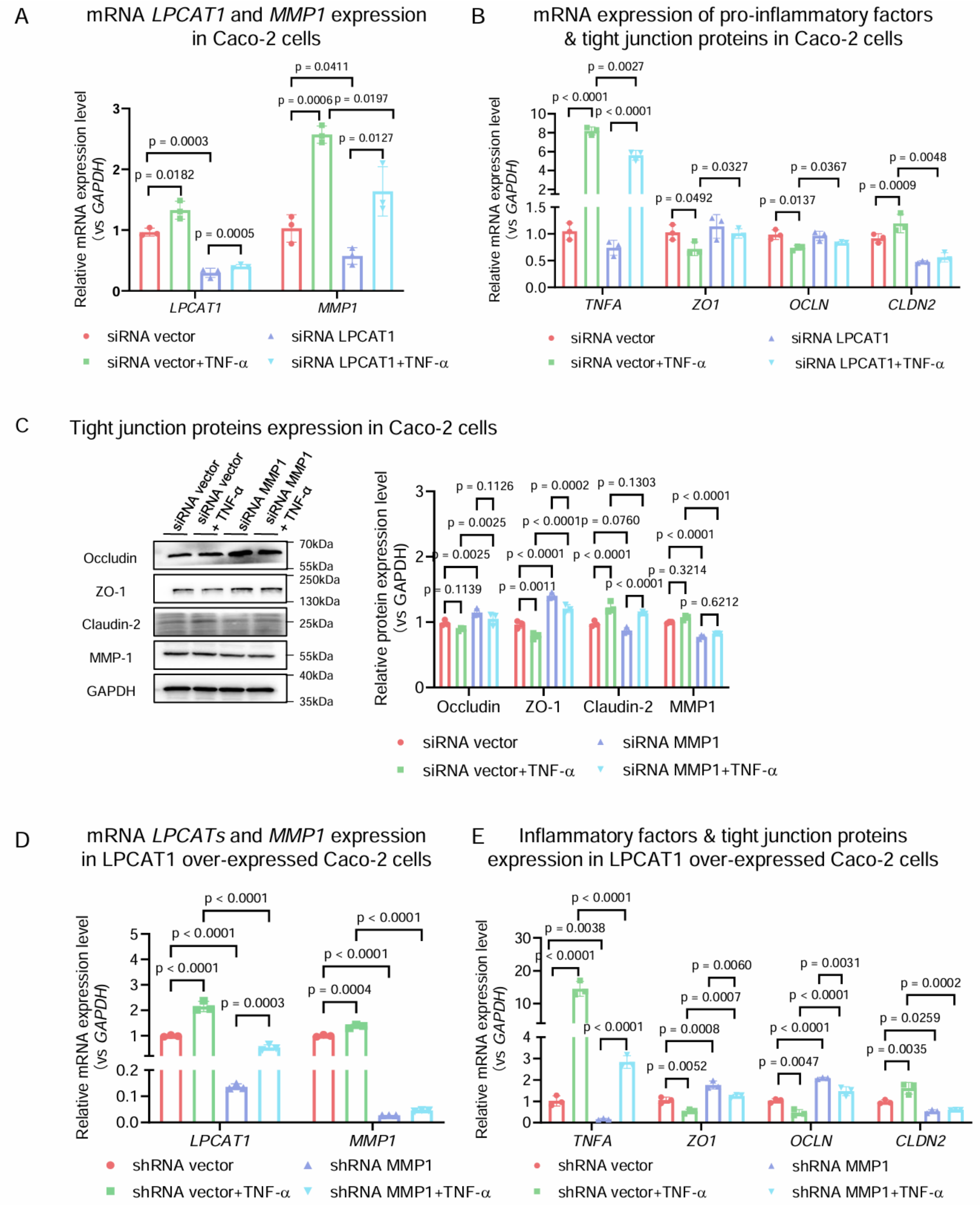


**Figure.S7 Inhibiting LPCAT1-MMP1 by genetic approaches restores intestinal barrier integrity and inflammatory responses in Caco-2 cells. (A-B)** mRNA expression levels of *LPCAT1*, *MMP1*, pro-inflammatory factors and tight junction proteins in WT and LPCAT1 knock down Caco-2 cells using LPCAT1 siRNA in the treatment of TNF-α (n=3 per group). **(C)** Western blot analysis and semi-quantification of ZO-1, Occludin and Claudin 2 in WT and MMP1 knock down Caco-2 cells using MMP1 siRNA in the treatment of TNF-α. **(D-E)** mRNA expression levels of *LPCAT1*, *MMP1*, pro-inflammatory factors, and tight junction proteins in LPCAT1 over-expressed Caco-2 cells and MMP1 knockout LPCAT1 over-expressed Caco-2 cells using MMP1 shRNA in the treatment of TNF-α (n=3 per group). Data were analyzed using a two-tailed one-way ANOVA.

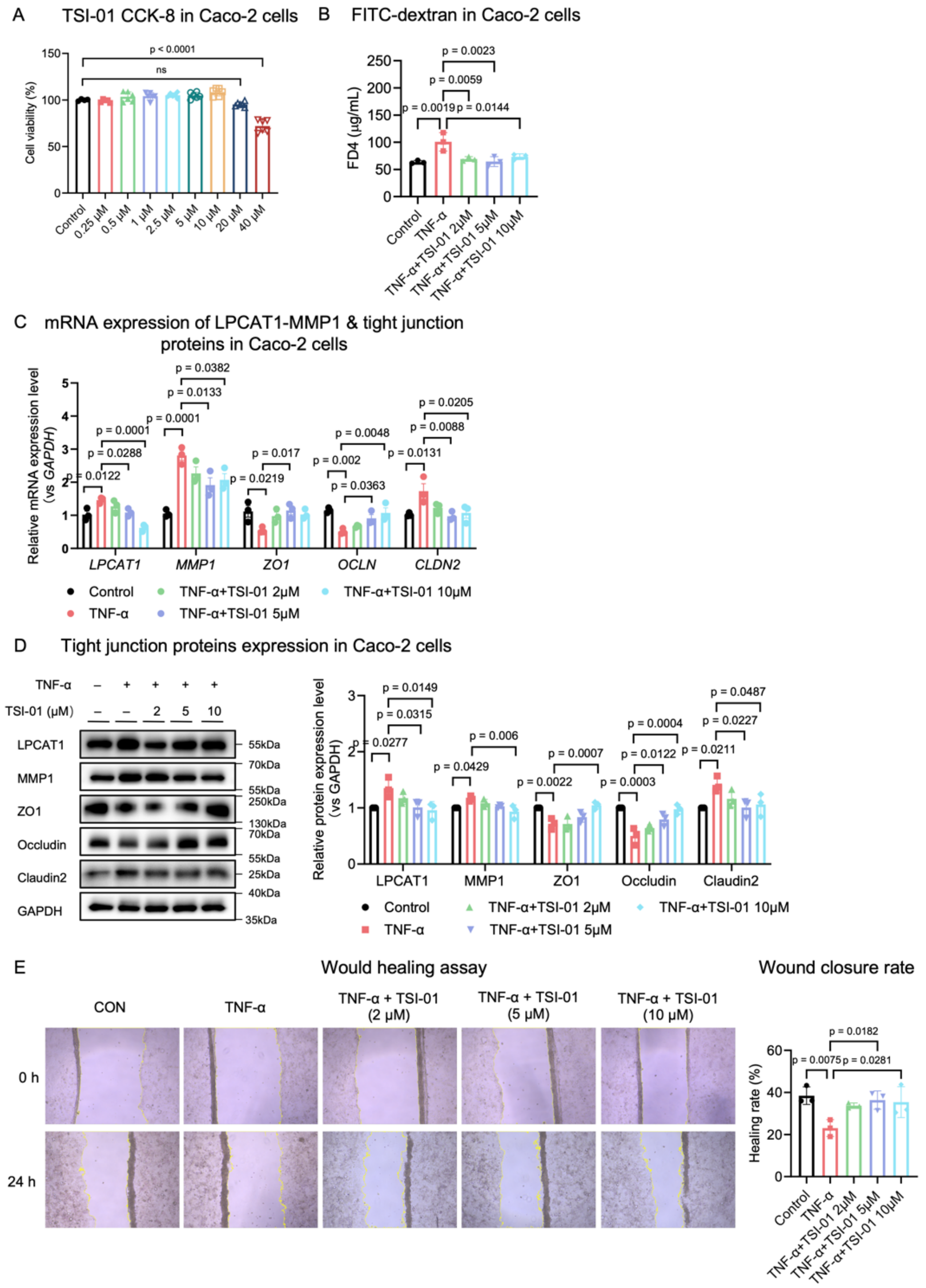


**Figure.S8 Inhibiting LPCAT1-MMP1 by an LPCAT1 inhibitor TSI-01 restores intestinal barrier integrity and inflammatory responses in Caco-2 cells. (A)** Cell viability of caco-2 cells treated with various concentrations of TSI-01 (n=6 per group). **(K)** FITC-Dextran of Caco-2 cells monolayer model treated with various concentrations of TSI-01 under TNF-α treatment (n=3 per group). **(C)** mRNA expression levels of pro-inflammatory factors and tight junction proteins in Caco-2 cells treated with various concentrations of TSI-01 under TNF-α treatment (n=3 per group). **(D)** Western blot analysis and semi-quantification of LPCAT1, MMP1, ZO-1, Occludin and Claudin 2 in Caco-2 cells treated with various concentrations of TSI-01 under TNF-α treatment (n=3 per group). **(E)** Wound healing closure rate in Caco-2 cells treated with various concentrations of TSI-01 under TNF-α treatment (n=3 per group). Data were analyzed using a two-tailed one-way ANOVA.

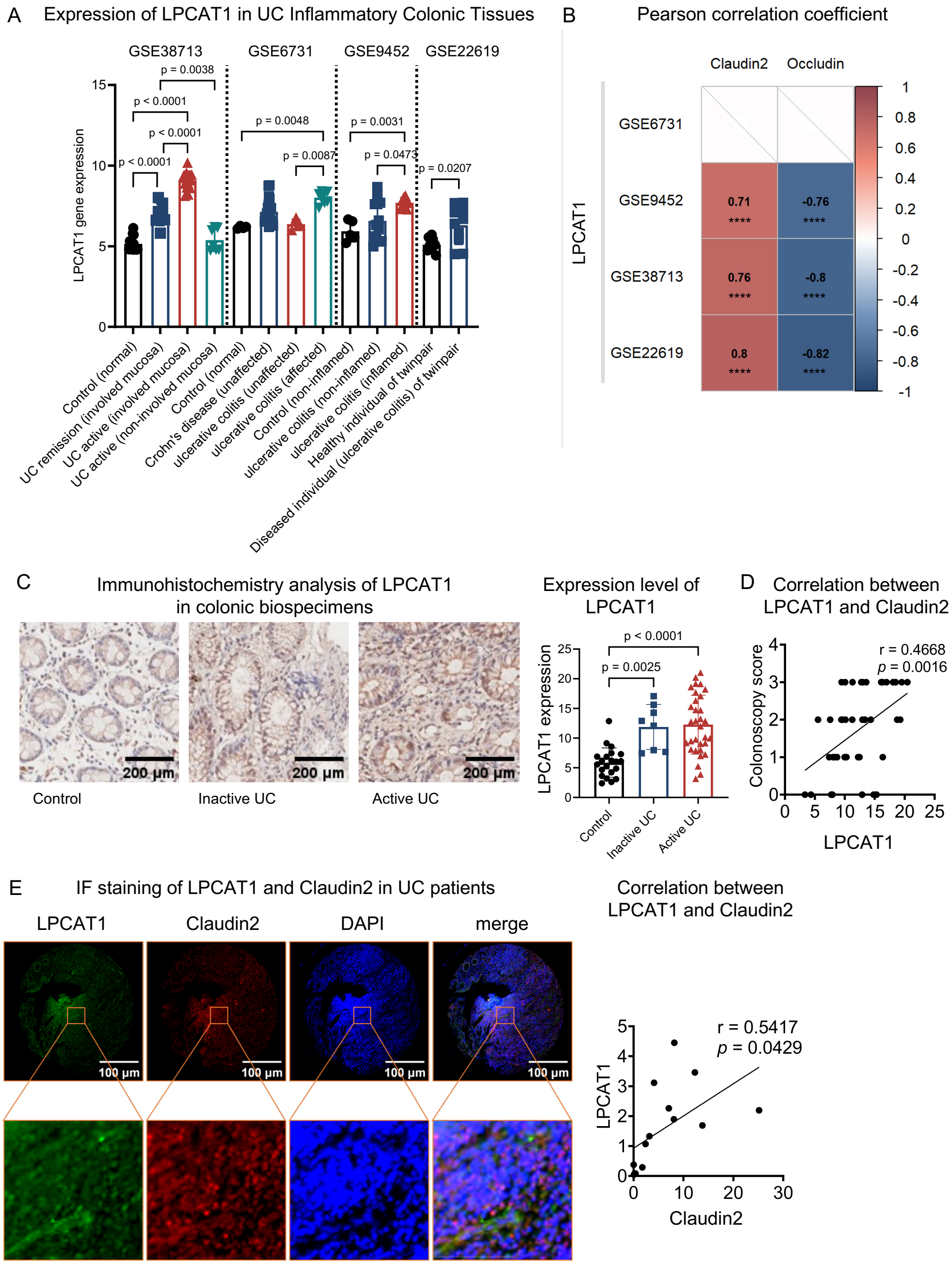


**Figure.S9 LPCAT1 is positively associated with intestinal barrier dysfunction in patients with UC. (A)** LPCAT1 expression levels in colonic tissues from patients with UC based on the GEO database. **(B)** Pearson correlation analysis between LPCAT1 expression and the levels of Claudin2 and Occludin in colonic tissues from patients with UC. **(C)** Immunohistochemistry analysis of LPCAT1 expression in colonic tissues from UC patients with both inactive and active stages and marginal normal tissues from patients with colon tumors. **(D)** Non-parametric spearman correlation analysis between LPCAT1 expression and colonoscopy scores in colonic tissues from UC patients with both inactive and active stages. **(E)** IF staining and non-parametric spearman of LPCAT1 and Claudin2 in inflamed colonic tissues of UC patients.
