## Supplemental Table 1 for "Gut microbiota-derived lithocholic acid leads to intestinal barrier dysfunction via LPCAT1 in irritable bowel syndrome"

Table.S1 Reagents and resource details in this study

| Reagent or Resource | SOURCE | IDENTIFIER |
|---|---|---|
| **Primers** | | |
| H_LPCAT1-F | 5'-CACAACCAAGTGGAAATCGAG-3' | DOI: 10.1186/s13046-017-0525-1 |
| H_LPCAT1-R | 5'-GCACGTTGCTGGCATACA-3' | |
| H_LPCAT2-F | 5'-AAAGCGAACAACATCAGGAG-3' | |
| H_LPCAT2-R | 5'-GAGCTGGCAGAAAGTAAGCA-3' | |
| H_LPCAT3-F | 5'-ATCACTGCCGTCCTCACTAC-3' | |
| H_LPCAT3-R | 5'-AGTCAACAGCCAAACCAATC-3' | |
| H_LPCAT4-F | 5'-TTGTGGATGTGGAGTTCCTT-3' | |
| H_LPCAT4-R | 5'-ACTTTTCCCAGTTCCCAGAG-3' | |
| H_TNF-α-F | 5'-AAGGACACCATGAGCACTGAAAGC-3' | Sangon Biotech (Shanghai) Co., Ltd. |
| H_TNF-α-R | 5'-AGGAAGGAGAAGAGGCTGAGGAAC-3' | |
| H_IL-6-F | 5'-GACAGCCACTCACCTCTTCAGAAC-3' | |
| H_IL-6-R | 5'-GCCTCTTTGCTGCTTTCACACATG-3' | |
| H_ZO1-F | 5'-CAGGAGGCATTGCTGATGAT-3' | |
| H_ZO1-R | 5'-GAAGGCTGGGGCTCATTT-3' | |
| H_Occludin-F | 5'-ACTTCGCCTGTGGATGACTTCAG-3' | |
| H_Occludin-R | 5'-TTCTCTTTGACCTTCCTGCTCTTCC-3' | |
| H_Claudin2-F | 5'- AGGTGCTGCTGAGGATAGACTGAC-3' | |
| H_Claudin2-R | 5'-TGGCGAGCATCCTTGGCAATTC-3' | |
| H_MMP1-F | 5'-TTACACGCCAGATTTGCCAAGAG-3' | |
| H_MMP1-R | 5'-TCAGAGGTGTGACATTACTCCAGAG-3' | |
| H_Beta-actin-F | 5'-CACCATTGGCAATGAGCGGTTC-3' | |
| H_Beta-actin-R | 5'-AGGTCTTTGCGGATGTCCACGT-3' | |
| H_GAPDH-F | 5'-ACCCACTCCTCCACCTTTGAC-3' | |
| H_GAPDH-R | 5'-TCCACCACCCTGTTGCTGTAG-3' | |
| M_LPCAT1-F | 5'-CCTGTATGCCAGCAATGTGAGG-3' | OriGene CAT#: MP207687 |
| M_LPCAT1-R | 5'-TCAGCAGGCAAGCGAAGTTGTC-3' | |
| M_LPCAT2-F | 5'-CTTCGCTGGTATCACGGAATGAG-3' | OriGene CAT#: MP207688 |
| M_LPCAT2-R | 5'-CCATTCTCCACCTGATGTCGCT-3' | |
| M_LPCAT3-F | 5'-CCATCTCTTCCACACCTTCACG-3' | OriGene CAT#: MP208006 |
| M_LPCAT3-R | 5'-GGATGAGGAACTGAAGCACGAC-3' | |
| M_LPCAT4-F | 5'-CGTGCATGAGTTACACCTCTCC-3' | OriGene CAT#: MP200007 |
| M_LPCAT4-R | 5'-CAGTGAGACCTGCTACTTGGAG-3' | |
| M_IL-6-F | 5'-TGTAACTGGCCTGCAGTAGC-3' | Self-design |
| M_IL-6-R | 5'-CTTTCCCTCACCCTAGCAGC-3' | |
| M_TNF-a-F | 5'-ATAGCAAATCGGCTGACGGT-3' | doi: 10.1074/jbc.M114.634337 |
| M_TNF-a-R | 5'-AGCCGATGGGTTGTACCTTG-3' | |
| M_iNOs-F | 5'-CAGAAGGGGACTGGGTTGTC-3' | doi.org/10.1002/jsfa.12477 |
| M_iNOs-R | 5'-GTGGCCATAAGCCAGACTGT-3' | |
| M_ZO1-F | 5'-ACCCGAAACTGATGCTGTGGATAG-3' | Sangon Biotech (Shanghai) Co., Ltd. |
| M_ZO1-R | 5'-GCTGGCTGGCTGTACTGTGAG-3' | |
| M_MMP1-F | 5'-GACGAGAGTGGTGGTGGTGAG-3' | |
| M_MMP1-R | 5'-CTGGATGGTGGCTAAGGTCTGG-3' | |
| M_Claudin2-F | 5'-AGCATTGTGACGGCGGTTGG-3' | |
| M_Claudin2-R | 5'-GGCAGCCTGGATGTCAGCAG-3' | |
| M_Occludin-F | 5'-AGGCAGCCTCGGTACAGCAG-3' | |
| M_Occludin-R | 5'-AGGCAGCCTCGGTACAGCAG-3' | |
| M_beta-actin-F | 5'-GGCTGTATTCCCCTCCATCG-3' | |
| M_beta-actin-R | 5'-CCAGTTGGTAACAATGCCATGT-3' | |
| M_GAPDH-F | 5'-ACGGCAAATTCAACGGCACAG-3' | |
| M_GAPDH-R | 5'-ACACCAGTAGACTCCACGACATAC-3' | |
| **Antibodies** | | |
| Rabbit anti-LPCAT1 Polyclonal Antibody(50μL) | Absin | abs116528 |
| MMP-1(E9S9N) Rabbit mAb #54376 | Cell Signalling Technology | 54376S |
| LPCAT1 (E4V4B) Rabbit mAb | Cell Signalling Technology | 57411S |
| GAPDH (14C10) Rabbit mAb | Cell Signaling Technology | 2118S |
| β-Actin (8H10D10) Mouse mAb (HRP Conjugate), 100ul | Cell Signaling Technology | 12262S |
| MMP1 Polyclonal Antibody (100μL) | Proteintech | 10371-2-AP |
| Anti-ZO1 tight junction Rabbit mAb | Abcam | ab96587 |
| Anti-Occludin [EPR20992] Rabbit mAb | Abcam | ab216327 |
| Claudin-2 (E1H9O) Rabbit mAb #48120 | Cell Signaling Technology | 48120S |
| **Chemicals** | | |
| Cholic acid-d4 | Sigma-Aldrich | 330256W |
| Deoxycholic acid-d4 | Sigma-Aldrich | 330257W |
| Isoallolithocholic acid(LCA) | Sigma-Aldrich | L6250 |
| Cholic acid (CA) | Sigma-Aldrich | C1129 |
| Sodium chenodeoxycholate (CDCA) | Sigma-Aldrich | C8261 |
| hyodeoxycholic acid (HDCA) | Sigma-Aldrich | H3878 |
| Sodium deoxycholate (DCA) | Sigma-Aldrich | D6750 |
| Isoallolithocholic acid (LCA) | Sigma-Aldrich | L6250 |
| Ursodeoxycholic acid (UDCA) | Sigma-Aldrich | U5127 |
| hyocholic acid (HCA) | Sigma-Aldrich | 700159P |
| iso-LCA | Sigma-Aldrich | 700195P |
| iso-DCA | Cayman Chemical | 29890 |
| Sodium taurocholate hydrate (TCA) | Sigma-Aldrich | 86339 |
| Taurochenodeoxycholic acid (TCDCA) | Sigma-Aldrich | 700249P |
| Taurohyocholic acid (THCA) | Cayman Chemical | 22669 |
| Sodium taurohyodeoxycholate hydrate (THDCA) | Sigma-Aldrich | T0682 |
| Sodium tauroursodeoxycholate (TUDCA) | Cayman Chemical | 9003379 |
| Sodium taurodeoxycholate hydrate (TDCA) | Sigma-Aldrich | T0875 |
| Taurolithocholic acid (TLCA) | Sigma-Aldrich | 700252P |
| Glycocholic acid hydrate (GCA) | Sigma-Aldrich | G2878 |
| Glycochenodeoxycholic acid (GCDCA) | Sigma-Aldrich | 700266P |
| Glycohyocholic acid (GHCA) | Cayman Chemical | 22670 |
| Glycoursodeoxycholic acid (GUDCA) | Sigma-Aldrich | 06863 |
| Glycohyodeoxycholic Acid(GHDCA) | Macklin | G922890 |
| Sodium glycodeoxycholate (GDCA) | Sigma-Aldrich | G9910 |
| Glycolithocholic acid (GLCA) | Sigma-Aldrich | 700268P |
| 1.5 M Tris-HCl Buffer (pH = 8) | Solarbio | T1010 |
| 1 M Tris-HCl Buffer (pH = 6.8) | Solarbio | T1020 |
| RIPA lysis buffer | Solarbio | R0020 |
| RNAisoPlus (Takara). | Takara | 9108 |
| Pierrce$^{TM}$BCA Protein Assay Reagent A | Thermo Scientific | Y1377216 |

| | | |
|---|---|---|
| Pierrce$^{TM}$BCA Protein Assay Reagent B | Thermo Scientific | YJ375831 |
| TEMED | BIO-RAD | 161-0801 |
| PrimeScript RT Master Mix | Takara | RR036A |
| PowerTrack™SYBR Green Master Mix | ThermoFisher | A46109 |
| Phosphate-buffered saline (PBS) | Gibco | 18912-014 |
| Human Lysophosphatidylcholine Acyltransferase 1 (LPCAT1) Protein | Abbexa | abx168194 |
| Protein Labeling Kit RED-NHS 2nd Generation | NanoTemper | MO-L011 |
| Cycloheximide | MedChemExpress | HY-12320 |
| **Biological samples** | | |
| Mice serum | C57BL/6J male mice | NA |
| Mice feces | C57BL/6J male mice | NA |
| Human serum | Clinical Trials | NA |
| Human feces | Clinical Trials | NA |
| **Cell line** | | |
| Caco-2 cell | American Type Culture Collection (ATCC) | HTB-37 |
| **Human study** | | |
| Human FABP2 ELISA Kit | Abcam | ab193700 |
| Human LBP ELISA Kit | Abcam | ab279407 |
| **Experimental models** | | |
| C57BL/6J male mice | NA | NA |
| Rosa26-LSL-LPCAT1$^{flox/-}$ C57BL/6J male mice | Cyagen Biosciences (Guangzhou) | NA |
| villin-Cre LSL-LPCAT1$^{flox/flox}$C57BL/6J male mice | Cyagen Biosciences (Guangzhou) | NA |
| **Animal study** | | |
| Dextran Sulfate Sodium Salt (DSS) - Colitis Grade (36,000-50,000 MW) | MP Biomedicals | 9011-18-1 |
| Isoflurane | Piramal | 330250 |
| Hematoxylin-Eosin(HE) Stain Kit | Solarbio | G1120 |
| Nuclear Fast Red Soulution | Abcam | ab150662 |
| Alcian Blue Ph2.5 | Abcam | 56735 |
| Fluorescein isothiocyanate–dextran average mol wt 3,000-5,000 | Sigma-Aldrich | FD4-1G |
| Mouse FABP2/I-FABP ELISA Kit | BIO THCHNE | NBP2-82214 |
| Mouse Matrix Metalloproteinase 1 (MMP1) ELISA Kit | JiangLai Biotech | JL12602-96T |
| Mouse TNF-α ELISA Kit | JiangLai Biology | JL10484 |
| Mouse IL-6 ELISA Kit | JiangLai Biology | JL12602-96T |
| **Software and algorithms** | | |
| Prism 10 | GraphPad | Home - GraphPad |
| Imgae J | Maryland | ImageJ |
| KneadData | github.com/biobakery/kneaddata/ | DOI: 10.1186/s13059-019-1891-0 |
| MetaPhlAn3 version 3.0.5 | MetaPhlAn | DOI: 10.7554/elife.65088 |
| R Statistical Software 3.4.4 | R Core Team | DOI: 10.1080/00031305.2017.1375986 |
| Cytoscape v3.9.0 | Cytoscape | Cytoscape 3.9.0 Release Notes |
| MO.Affinity Analysis Software | NanoTemper | |
| PyMOL 3.1 | PyMOL | PyMOL by Schrödinger |
| **Others** | | |
| 3mm steel grinding beads | Servicebio | G103 |
| 4mm steel grinding beads | Servicebio | G104 |
| TissueLyser II | QIAGEN | NA |
| UPLC column C18 (2.1x100mm 1.7μm) | Waters | NA |
| R500 Small Animal Anesthesia Machine | RWD | NA |
| EasyMount/VCC MC6plus | JINGONGHONGTAI | NA |
| UC_2M Ussing Chamber System | JINGONGHONGTAI | NA |
| C4300 Electrode Vertical Insertion Ussing Chamber | JINGONGHONGTAI | NA |
| DA1008 Data Acquisition Unit | JINGONGHONGTAI | NA |
| EKTip-50V | JINGONGHONGTAI | NA |
| EKTip-50C | JINGONGHONGTAI | NA |
| EKVC | JINGONGHONGTAI | NA |
| Millicell® ERS-2 Voltohmmeter | MERCK | NA |
| Invitrogen Countess 3 FL | Thermo Fisher Scientific | NA |
| Spectra Max iD5 | Molecular Devices | NA |
| Nano Drop One | Thermo Fisher Scientific | NA |
| Veritipro 96 well Thermo Cycle | Thermo Fisher Scientific | NA |
| Quant Studio 7 Flex | Thermo Fisher Scientific | NA |
| PowerPac Basic | BIO-RAD | NA |
| ChemiDoc MP imaging system | BIO-RAD | NA |
| Fully Automated Fluorescense Upright Microscope | Leica | NA |
| THUNDDER Imager 3D Tissue | Leica | NA |
| Cryostar NX70 | Thermo Fisher Scientific | NA |
| Monolith X | NanoTemper | NA |
